# In Vivo Blood Kinetics and Transcript Integrity of Three mRNA–Lipid Nanoparticle Vaccines in Humans

**DOI:** 10.64898/2026.03.13.26348310

**Authors:** Stephen J. Kent, Shiyao Li, Thakshila H. Amarasena, Arnold Reynaldi, Michael G. Leeming, Jennifer A. Juno, Adam K. Wheatley, Georgia Deliyannis, Dale I. Godfrey, Terry Nolan, Colin W. Pouton, Miles P. Davenport, Yi Ju

## Abstract

mRNA–lipid nanoparticle (LNP) vaccines are detectable in human blood after vaccination, but platform-specific differences in systemic persistence and transcript integrity remain poorly defined. We analyzed serial blood samples from 73 participants receiving Moderna mRNA-1273 (three formulations), Pfizer/BioNTech BNT162b2, or an investigational receptor-binding domain (RBD) mRNA vaccine (three different doses). Using droplet digital polymerase chain reaction (ddPCR) assays, we quantified total and long-range linked (“intact”) vaccine mRNA, and we measured vaccine-specific ionizable lipids by liquid chromatography–mass spectrometry (LC–MS). Across platforms, mRNA decay was fastest for mRNA-1273, intermediate for BNT162b2, and slowest for the RBD vaccine, with ionizable lipid decay following the same rank order. Notably, intact spike mRNA declined two-fold faster after mRNA-1273 than BNT162b2 vaccination. Kinetics modelling revealed platform-dependent coupling of mRNA and lipid kinetics: intact mRNA tracked closely with SM-102 for mRNA-1273, whereas ALC-0315 persisted longer than intact mRNA for BNT162b2. A ten-fragment linkage ddPCR panel spanning the spike transcript showed lower linkage toward 3′-proximal regions that mirrored the administered mRNA-1273 formulation. Together, these data establish a quantitative framework for benchmarking mRNA–LNP platform kinetics and transcript integrity in humans.

## Introduction

mRNA vaccines and therapeutics are transforming modern medicine, yet the *in vivo* distribution, persistence, and degradation of mRNA–lipid nanoparticle (LNP) formulations remain incompletely understood^1^. The two most widely used SARS-CoV-2 mRNA vaccines are serially updated versions of the Moderna mRNA-1273 (Spikevax) and Pfizer/BioNTech BNT162b2 (Comirnaty) (hereafter referred to as “Moderna” and “Pfizer”, respectively). In parallel, multiple next-generation mRNA vaccines are in development and clinical testing, including candidates encoding shorter spike segments such as the receptor-binding domain (RBD)^2^. Notably, Moderna has generally shown modestly higher immunogenicity and effectiveness than Pfizer, but is administered at a higher dose (currently 50 µg vs 30 µg) and is associated with greater reactogenicity^3,4^. The molecular and biophysical factors contributing to these platform-level differences remain poorly defined.

Defining how vaccine components behave in human blood provides a practical window into *in vivo* kinetics of mRNA–LNPs that would otherwise require sampling of human tissues. We and others have shown that both vaccine mRNA and vaccine-specific ionizable lipids can be quantified in serial blood samples for weeks following vaccination^5–7^. These studies enabled detailed kinetic modeling and identified clinical and immunological correlates of circulating vaccine mRNA levels and decay^7^. Although anti-polyethylene glycol (PEG) antibodies were initially hypothesized to accelerate clearance, given that PEG-lipids are present in mRNA–LNPs, no significant association with faster decay was observed. Instead, higher peak concentrations of circulating vaccine mRNA and ionizable lipid correlated with stronger boosting of anti-PEG antibody levels^7^. However, a head-to-head comparison of mRNA decay, lipid persistence, and transcript integrity across different mRNA–LNP vaccine platforms in humans has not been reported.

Improving mRNA stability and limiting degradation could, in principle, enhance vaccine performance by increasing the duration and magnitude of antigen expression. In small animal models, increasing mRNA stability or reducing degradation can increase antigen expression and improve vaccine responses^8^. Beyond vaccines, systemically administered mRNA–LNP therapeutics have entered clinical trials, including candidates for *in vivo* gene editing and immunotherapy.^9–11^ Because intravenously administered mRNA–LNPs necessarily transit through and interact with blood before reaching target tissues, understanding the fate of LNP-formulated mRNA in human blood is increasingly important for improving the efficacy and safety of emerging mRNA–LNP therapeutics.

A key unresolved question is how LNP-formulated mRNA loses integrity *in vivo*. Naked mRNA is rapidly degraded by extracellular RNases^12^, whereas LNP encapsulation provides substantial protection. Nonetheless, several aspects of transcript integrity in human blood remain unclear: i) the extent to which integrity loss reflects fragmentation already present in the administered formulation versus post-vaccination *in vivo* degradation; (ii) how rapidly long-range mRNA integrity is lost in blood over time; and (iii) whether specific transcript regions are disproportionately vulnerable to degradation.

Here, we analyzed serial blood samples from participants receiving licensed Moderna or Pfizer spike mRNA vaccines or an investigational RBD mRNA–LNP vaccine (hereafter referred to as “mRNA–RBD”) from a phase I clinical trial (Fig. 1A)^2^. We compared the blood kinetics of total mRNA, long-range linked mRNA (as a measure of transcript integrity), vaccine-specific ionizable lipid, and anti-PEG and anti-spike antibodies across platforms (Fig. 1B). To map integrity loss along the transcript, we further developed a panel of linkage droplet digital polymerase chain reaction (ddPCR) assays spanning the spike coding region. Together, these data provide a comprehensive, platform-level view of mRNA and lipid persistence and transcript integrity in human blood, establishing a quantitative framework for benchmarking mRNA–LNP formulations *in vivo*.

**Figure 1.**
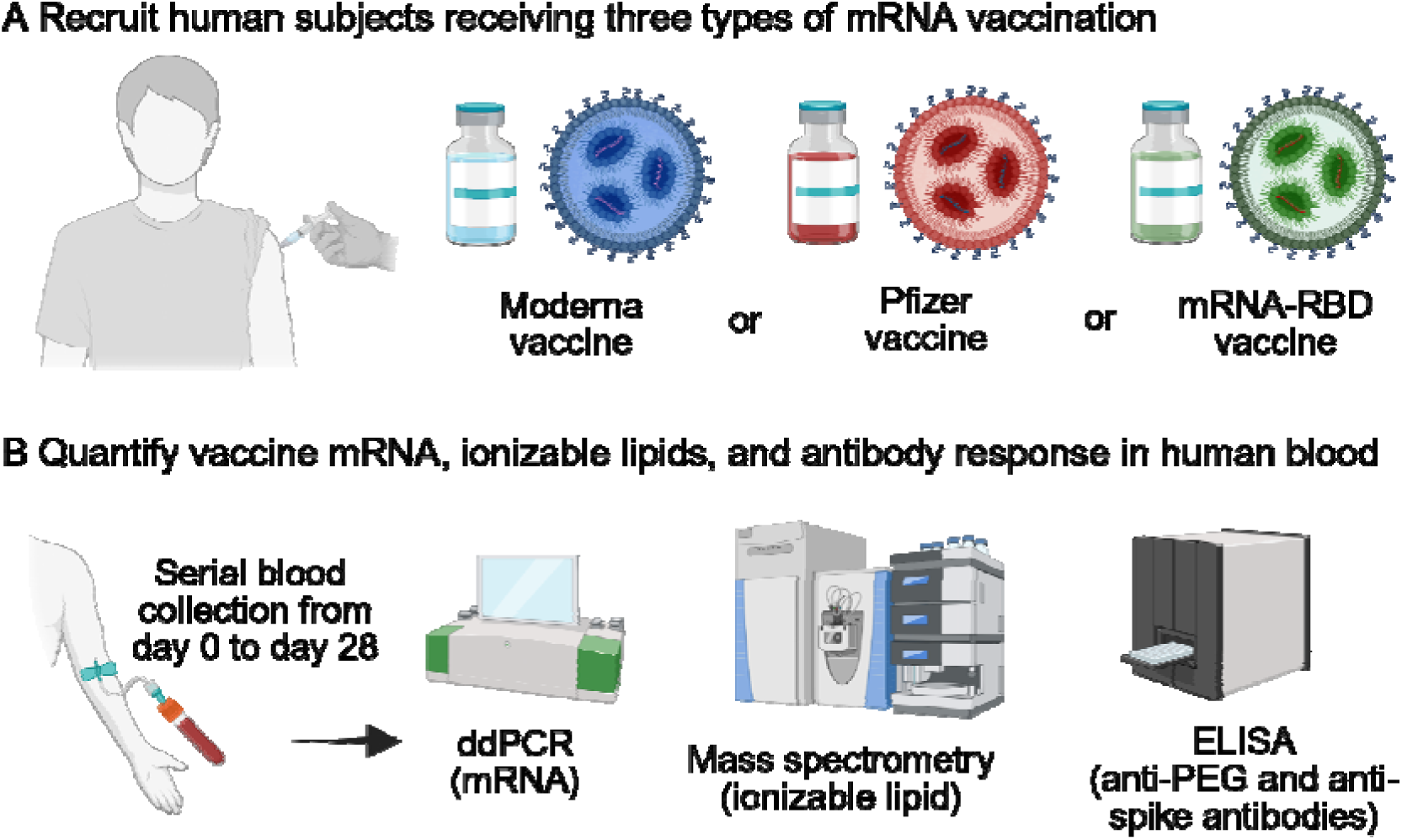
Schematic illustration of the workflow for comparative analysis of three mRNA vaccines in human blood. A) Three cohort of human subjects were recruited who received either Moderna, Pfizer, or mRNA-RBD vaccination. B) Serial blood samples were collected before and after vaccination and analyzed by ddPCR, mass spectrometry and ELISA to quantify vaccine mRNA, ionizable lipid, and antibody response (anti-PEG and anti-spike), respectively.

## Results

### SARS-CoV-2 mRNA–LNP vaccine clinical studies

We analyzed longitudinal blood samples from 73 participants intramuscularly vaccinated with Moderna mRNA-1273 (n = 29; mRNA dose = 50 µg; type = bivalent ancestral + BA.1, bivalent ancestral + BA.5, or monovalent XBB.1.5), Pfizer/BioNTech BNT162b2 (n = 12; mRNA dose = 30 µg; type = ancestral), or an investigational mRNA–RBD vaccine (n = 32; mRNA dose = 10, 20, or 50 µg; type = B.1.351 RBD) (details of participants and vaccination in Table S1). A total of 327 samples (median 3, range 2−15 samples per participant) were serially collected pre-vaccination (day 0) and from 4 h to day 28 post-vaccination or later. Sampling schedules differed across studies: Moderna and Pfizer cohorts included multiple early time points, whereas the mRNA–RBD trial sampled primarily on days 0, 7, and 28. Plasma (with EDTA or heparin anticoagulation) and serum samples were aliquoted and stored at −80 °C.

### Vaccine mRNA exhibits platform-specific decay kinetics

Using reverse transcription (RT)-ddPCR assays, vaccine mRNA was detectable in blood after vaccination and declined over time across all three platforms (Fig. 2A–C, Fig. S1A–C). All pre-vaccination samples were undetectable (0 copies µL^−1^) for vaccine mRNA. In the Moderna cohort, where early sampling was most frequent, total mRNA typically peaked at day 1–2 (median 1) post-vaccination at a peak concentration of 0.001 to 0.529 (median 0.095) ng mL^−1^ (Fig. 2A; Fig. S1A). At day 6–7 post-vaccination, circulating mRNA concentrations were not significantly different between Moderna and Pfizer (Fig. 2D), whereas mRNA–RBD levels were 35- and 109-fold lower than Moderna and Pfizer, respectively. Mixed-effects modeling of post-peak decay demonstrated platform-dependent decay rates (Fig. 2E), with Moderna exhibiting the fastest mRNA decline (median 0.389 dayL¹), Pfizer an intermediate decline (0.250 dayL¹), and mRNA–RBD the slowest (0.123 dayL¹).

**Figure 2.**
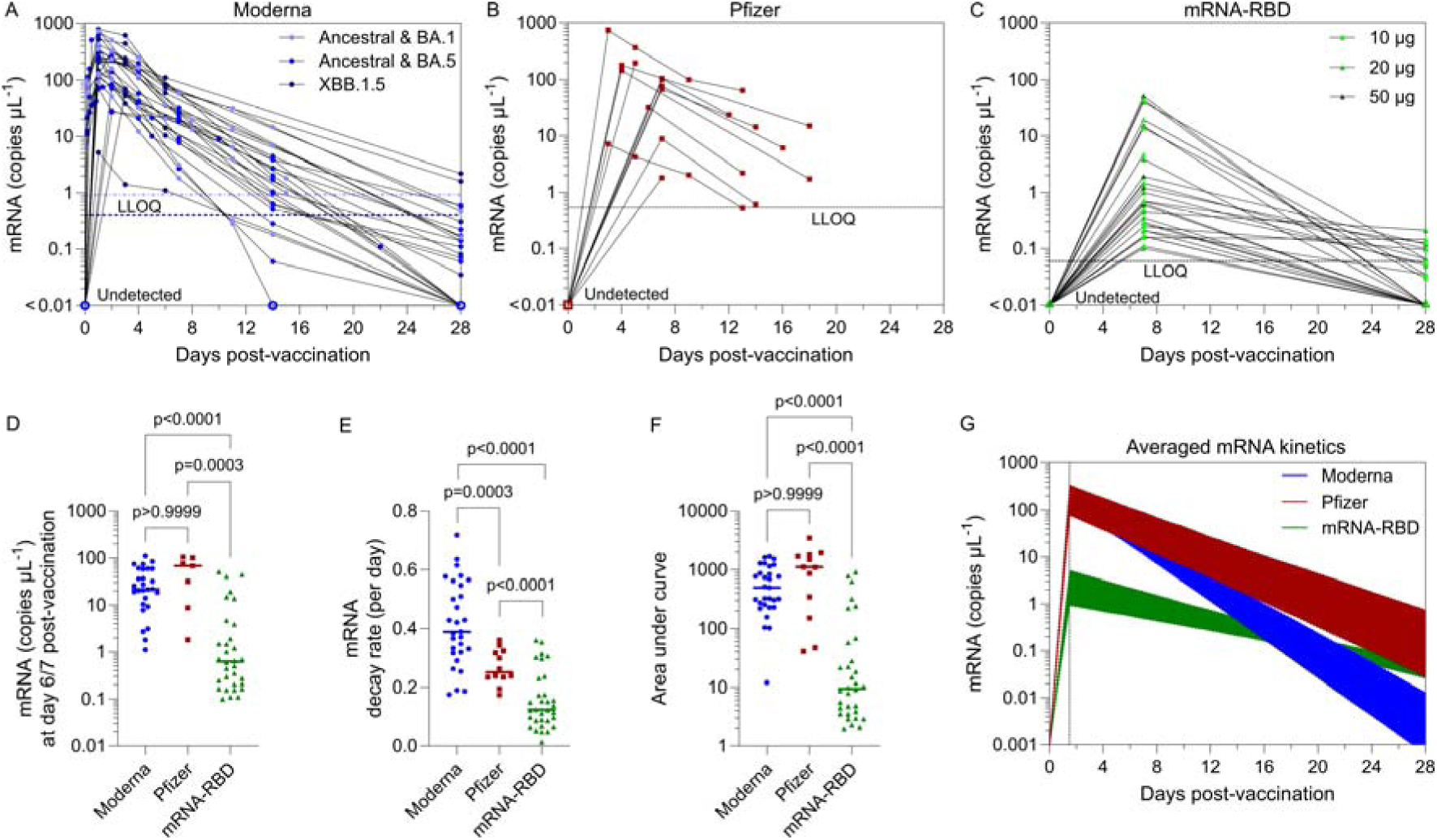
Comparison of *in vivo* vaccine mRNA kinetics in human blood following three types of SARS-CoV-2 mRNA vaccination (Moderna, Pfizer, or mRNA-RBD). (A–C) Longitudinal vaccine mRNA concentrations (copies µL^−1^) in human blood from seven cohorts receiving either (A) Moderna bivalent ancestral + BA.1, bivalent ancestral + BA.5, or monovalent XBB.1.5; (B) Pfizer; or (C) mRNA-RBD at 10, 20, or 50 µg doses. The lower limit of quantification (LLOQ; dashed line) was determined from linear standard curves (Figure S1D–G). In panel A, two the lower limits of quantifications (LLOQs) are shown: 0.4 copies µL^−1^ for Moderna XBB.1.5 (dark blue dashed line) and 0.93 copies µL^−1^ for Moderna bivalent vaccines (light blue dashed line). Undetected samples (0 copies μL^−1^) were plotted with open symbols. (D–G) Comparison of (D) mRNA concentration at day 6–7 post-vaccination, (E) post-peak mRNA decay rates, (F) post-peak area under the curve (AUC) of mRNA kinetics in blood, and (G) averaged mRNA kinetics across donors among the three vaccine types. In (D–F), each dot represents one participant, and the horizontal line indicates the median. In (G), averaged mRNA kinetics are shown as mean predictions from the best-fit linear model, with shaded regions indicating the 95% confidence interval bounds. Statistical analysis was performed using the nonparametric Kruskal–Wallis test with Dunn’s multiple comparisons in (D, F) and the likelihood ratio test in (E).

To enable cross-platform comparison of circulating mRNA exposure despite differences in sampling density, we assumed that circulating vaccine mRNA peaked at day 1–2 and calculated the post-peak area under the curve (AUC) from the fitted decay rates. Using this approach, post-peak mRNA exposure was not significantly different between Moderna and Pfizer (median AUC 498.4 vs 1104.7), whereas mRNA–RBD exposure was markedly lower (median AUC 9.4), corresponding to ∼53-fold and ∼117-fold lower exposure than Moderna and Pfizer, respectively (Fig. 2F,G). Within the Moderna cohort, post-peak AUC did not differ significantly among the BA.1- and BA.5-bivalent and XBB.1.5 formulations (Fig. S2A). Similarly, mRNA–RBD AUCs were not significantly different across the 10, 20, and 50 µg dose groups, with substantial inter-individual variability (Fig. S2D).

### Ionizable lipid exposure and clearance differ across vaccines

We next quantified each platform’s vaccine-specific ionizable lipid by targeted liquid chromatography–mass spectrometry (LC–MS) (SM-102 for Moderna, ALC-0315 for Pfizer, and Dlin-MC3-DMA for mRNA–RBD; LNP formulations in Table S2). All three ionizable lipids were negative (below the lower limit of quantification, LLOQ) in pre-vaccination samples but became readily detectable at early post-vaccination time points and declined thereafter (Fig. 3A–C, Fig. S3A–C). Analytical sensitivity and platform specificity were supported by lipid standard curves (Fig. S3D–G), representative chromatograms (Fig. S3H–K), and cross-platform negative controls (Fig. S4).

**Figure 3.**
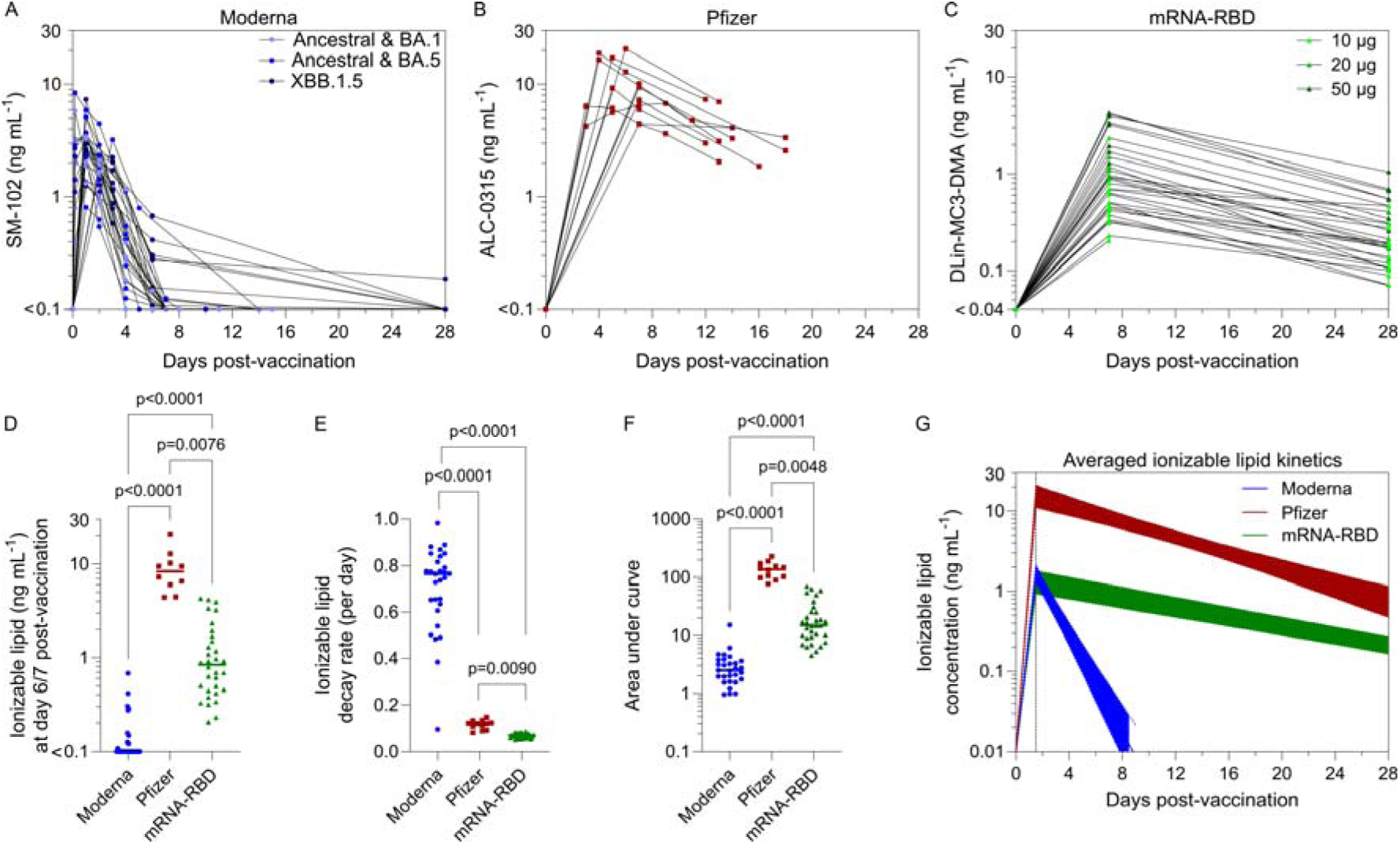
Comparison of *in vivo* ionizable lipid kinetics in human blood following Moderna, Pfizer, or mRNA-RBD vaccination. (A–C) Longitudinal ionizable lipid concentrations (ng mL^−1^) in human blood from seven cohorts who received either (A) Moderna bivalent ancestral + BA.1, bivalent ancestral + BA.5, or monovalent XBB.1.5 (formulated with SM-102); (B) Pfizer (formulated with ALC-0315); or (C) mRNA-RBD (formulated with Dlin-MC3-DMA) vaccination at 10, 20, or 50 µg doses. (D–G) Comparison of (D) ionizable lipid concentration at day 6–7 post-vaccination, (E) post-peak ionizable lipid decay rates, (F) post-peak AUC of ionizable lipid kinetics in blood, and (G) averaged ionizable lipid kinetics across donors among the three vaccine types. In (D–F), each dot represents one participant, and the horizontal line indicates the median. In (G), averaged ionizable lipid kinetics are shown as mean predictions from the best-fit linear model, with shaded regions indicating the 95% confidence interval bounds. Statistical analysis was performed using the nonparametric Kruskal–Wallis test with Dunn’s multiple comparisons in (D, F) and the likelihood ratio test in (E).

In the Moderna cohort, SM-102 peaked at 4 h to day 2 post-vaccination at a peak concentration of 0.81 to 8.40 (median 2.63) ng mL^−1^ (Fig. 3A). In the Pfizer cohort, the earliest post-vaccination time points were day 3–5, with ALC-0315 concentrations peaking at 4.24 to 18.89 (median 12.90) ng mL^−1^ (Fig. 3B). At day 6–7 post-vaccination, circulating ALC-0315 concentrations (median 8.33 ng mLL¹) were highest in the Pfizer cohort, followed by Dlin-MC3-DMA (median 0.84 ng mLL¹) in the mRNA–RBD cohort, whereas SM-102 in the Moderna cohort was often below the LLOQ (0.1 ng mLL¹) (Fig. 3D).

Consistent with the rank order observed for vaccine mRNA decay, lipid decay was fastest for Moderna (median 0.765 dayL¹), intermediate for Pfizer (median 0.120 dayL¹), and slowest for mRNA–RBD (median 0.069 dayL¹) (Fig. 3E). To compare overall circulating exposure across platforms, ALC-0315 lipid exposure was greatest in the Pfizer cohort (median AUC 136.5), corresponding to a 54.6- and 9.2-fold higher exposure than SM-102 in the Moderna cohort (median AUC 2.5) and Dlin-MC3-DMA in the mRNA–RBD cohort (median AUC 14.9), respectively (Fig. 3F,G). Within the Moderna cohort, SM-102 exposure was slightly higher after the XBB.1.5 booster (median AUC 4.0) than after the bivalent boosters (median AUC 1.9 for ancestral + BA.1 and 2.1 for ancestral + BA.5) (Fig. S2B). Within the mRNA–RBD trial, Dlin-MC3-DMA exposure increased with dose, with median AUCs of 9.1, 15.8, and 51.8 for 10, 20, and 50 µg, respectively (Fig. S2E).

### Integrity of circulating vaccine mRNA decreases faster after Moderna than Pfizer vaccination

The total vaccine mRNA measurements above rely on short amplicons (113 bp for Moderna and Pfizer; 134 bp for mRNA–RBD; Tables S3, S4) and therefore cannot distinguish long transcripts from shorter degraded fragments. To quantify long-range transcript integrity, we applied a linkage RT-ddPCR assay in which two widely separated targets on the same spike transcript are detected within individual droplets. An excess of double-positive droplets above that expected from random co-occupancy was used to infer the fraction of molecules that remain linked across the intervening region (hereafter termed “intact” for simplicity, noting this reflects integrity across the assayed span rather than full-length transcript integrity). The linkage assays spanned 1,599 bp for Moderna and 1,404 bp for Pfizer (Tables S5, S6), corresponding to ∼42% and ∼37%, respectively, of the 3,819-bp spike coding sequence. Because circulating mRNA–RBD vaccine mRNA levels were low at the earliest available time point (day 7), intact mRNA could not be reliably quantified for the mRNA–RBD cohort.

Across early post-vaccination samples, the fraction of intact spike mRNA was broadly similar between Moderna and Pfizer, generally ∼10–25% of total vaccine mRNA detected (Fig. 4A,B). In the Moderna cohort, the intact fraction was highest at 4 h to day 1 post-vaccination (7.4–19.7%, median 13.7%) and then declined over time (Fig. 4A). In the Pfizer cohort, sampling began at day 3–4, when the intact fraction was 12.3–26.8% (median 24.2%), and it subsequently decreased (Fig. 4B). Combining total vaccine mRNA concentrations with intact fractions, we calculated intact mRNA concentrations, which showed log-linear decay kinetics (Fig. 4C,D). By day 6–7 post-vaccination (among participants with a day 6–7 sample available), intact mRNA remained above the LLOQ in 20 of 27 Moderna participants (74%) and 6 of 7 Pfizer participants (86%), with median intact mRNA concentration of 2.1 and 8.3 copies µL^−1^, respectively. In Pfizer cohort, two additional participants without a day 6–7 sample had intact mRNA above the LLOQ at day 9 or day 18, bringing the overall proportion with detectable intact mRNA to 8 of 9 (89%).

**Figure 4.**
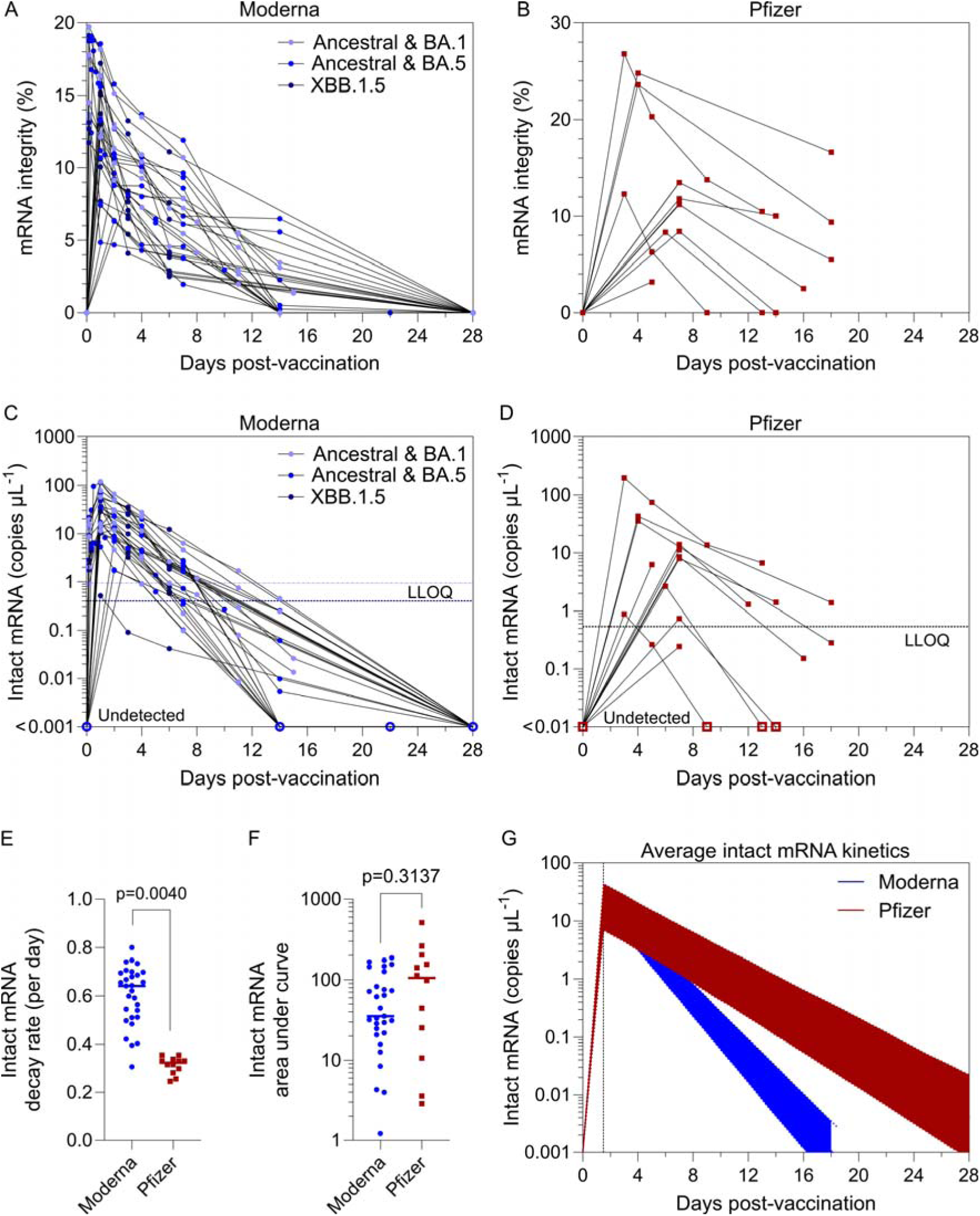
Comparison of *in vivo* vaccine mRNA integrity in human blood between Moderna and Pfizer vaccines. (A,B) Longitudinal vaccine mRNA integrity in human blood from four cohorts who received either (A) Moderna bivalent ancestral + BA.1, bivalent ancestral + BA.5, or monovalent XBB.1.5 or (B) Pfizer vaccination. (C,D) Longitudinal intact vaccine mRNA concentration (copies µL^−1^) in human blood from the four cohorts. The LLOQ (shown as a dashed line) was determined based on the linear standard curves of vaccine mRNA (Figure S1D–F). In panel C, two LLOQs are shown: 0.4 copies µL^−1^ for Moderna XBB.1.5 (dark blue dashed line) and 0.93 copies µL^−1^ for Moderna bivalent vaccines (light blue dashed line). Undetected samples (0 copies μL^−1^) were plotted with open symbols. (E–G) Comparison of (E) post-peak intact mRNA decay rates, (F) post-peak AUC of intact mRNA kinetics in blood, and (G) averaged intact mRNA kinetics across donors between the two vaccine types. In (E,F), each dot represents one participant, and the horizontal line indicates the median. In (G), averaged intact mRNA kinetics are shown as mean predictions from the best-fit linear model, with shaded regions indicating the 95% confidence interval bounds. Statistical analysis was performed using the nonparametric Mann−Whitney *U* test in (F) and the likelihood ratio test in (E).

Mixed-effects modeling of post-peak decay showed that intact mRNA declined ∼2-fold faster after Moderna than Pfizer vaccination (median 0.64 vs. 0.32 dayL¹; Fig. 4E). Consistent with these kinetics, modeled post-peak intact mRNA exposure (AUC) showed a higher median in the Moderna cohort than in the Pfizer cohort (median 105.4 vs 35.4), although this difference was not statistically significant due to substantial inter-individual variability (Fig. 4F,G). Intact mRNA exposure was similar across the three Moderna booster formulations (Fig. S2C).

### Platform-specific mRNA–ionizable lipid kinetic coupling

To enable direct comparison across analytes and vaccine platforms, we normalized each readout to its post-vaccination peak (set to 100%) and plotted best-fit decay trajectories for total mRNA, intact mRNA, and ionizable lipid (Fig. 5A), together with the corresponding half-life estimates (Fig. 5B). In the Moderna cohort, intact mRNA and SM-102 declined with closely aligned trajectories (blue dashed lines in Fig. 5A) and similar half-lives (Fig. 5B), consistent with our prior observations^7^, suggesting that circulating intact mRNA largely remains associated with ionizable lipid in blood. In contrast, in the Pfizer cohort, ALC-0315 decayed more slowly than intact mRNA (red dashed lines in Fig. 5A) and exhibited a longer half-life (Fig. 5B), indicating prolonged persistence of ionizable lipid in circulation, potentially within empty LNPs or partially degraded particles that no longer retain intact mRNA. In the mRNA–RBD cohort, intact mRNA could not be quantified reliably, but Dlin-MC3-DMA and total mRNA showed the slowest decay and longest half-lives among the three platforms (Fig. 5A,B).

**Figure 5.**
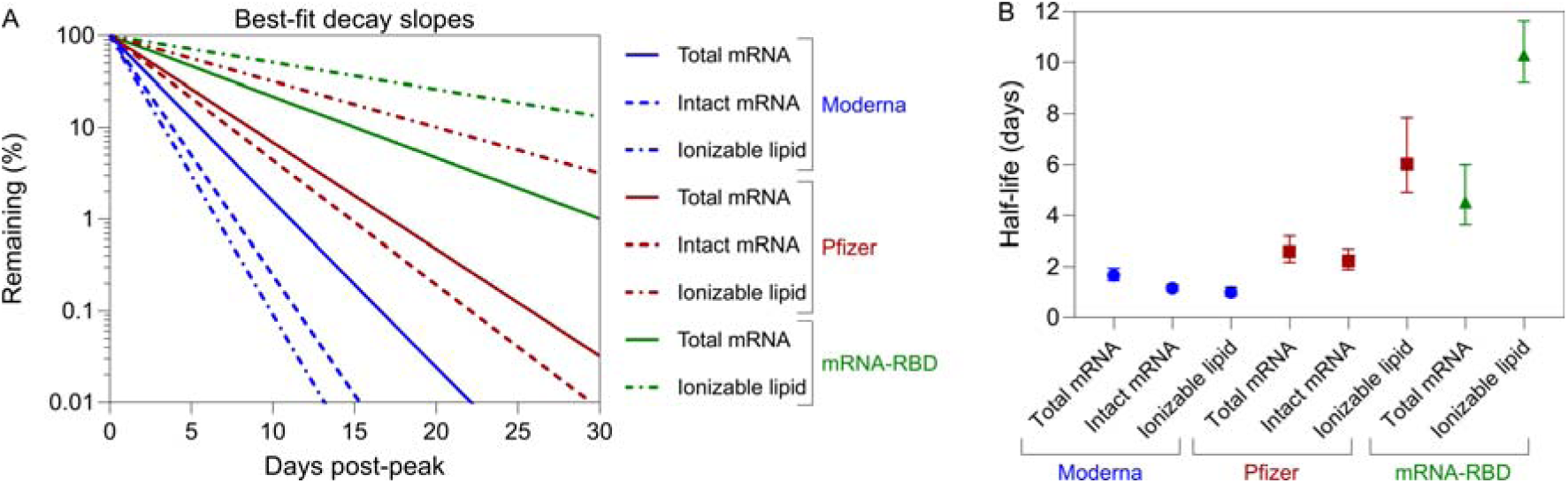
Comparison of *in vivo* decay kinetics of vaccine mRNA and ionizable lipids in human blood following Moderna, Pfizer, or mRNA-RBD vaccination. (A) Best-fit decay slopes of total mRNA, intact mRNA, and ionizable lipids across the three vaccines. The response at the peak time point for each parameter was normalized to 100%, and the percentage change over time illustrates the decline estimated from the best-fit linear model. (B) Half-life of total mRNA, intact mRNA, and ionizable lipids from the three vaccines, shown as the mean with upper and lower bound of 95% confidence intervals calculated across multiple donors in each cohort.

### Anti-PEG and anti-spike antibody responses across vaccine platforms

The PEG-lipid component of mRNA–LNP vaccines can induce anti-PEG antibodies. We and others have previously reported higher anti-PEG IgG and IgM responses following Moderna than Pfizer vaccination^13–15^. However, anti-PEG responses induced by experimental mRNA–LNP vaccines used in clinical trials, such as the mRNA–RBD vaccine studied here, remain unclear. We quantified plasma anti-PEG IgG and IgM titers by ELISA using our established method.^14^ Cohort immunization context differed: the Pfizer cohort received a prime (first-ever) SARS-CoV-2 mRNA vaccination, whereas the Moderna and mRNA–RBD cohorts received booster vaccinations in participants with prior COVID-19 mRNA–LNP vaccination (Table S1). Post-vaccination samples were collected at day 28 for the Moderna and mRNA–RBD cohorts, and between days 12–70 for the Pfizer cohort.

Prior to vaccination, anti-PEG IgG (end-point titer >1:10) was detectable in 24/29 (83%), 10/12 (83%), and 19/26 (73%) participants in the Moderna, Pfizer, and mRNA-RBD cohorts, respectively (Fig. 6A). Pre-existing anti-PEG IgM was detectable in 28/29 (97%), 7/12 (58%), and 11/26 (42%) participants in the Moderna, Pfizer, and mRNA–RBD cohorts, respectively (Fig. 6B). Following vaccination, anti-PEG antibody titers increased in a platform- and isotype-dependent manner. In the Moderna cohort, both anti-PEG IgG and IgM were boosted at day 28, with mean fold changes of 2.0 (range 1–6.7) and 5.9 (range 1–33.6), respectively (Fig. 6C,D). In contrast, after Pfizer vaccination, anti-PEG IgG showed no significant increase (mean fold change 1.1, range 1–1.4), whereas anti-PEG IgM increased significantly (mean fold change 1.6, range 1–5.1). In the mRNA–RBD cohort, more modest increases were observed, with mean fold changes of 1.2 (range 1–2.1) for anti-PEG IgG and 1.8 (range 1–11.9) for anti-PEG IgM. Overall, anti-PEG IgM showed a greater boost than anti-PEG IgG across the three vaccine platforms, and Moderna induced larger increases in both anti-PEG IgG and IgM than Pfizer or mRNA–RBD. Within the Moderna cohort, anti-PEG responses were similar across the three Moderna booster formulations (Fig. S5A,B), and no clear dose-dependent trend was observed across the mRNA–RBD dose groups (Fig. S5C,D).

**Figure 6.**
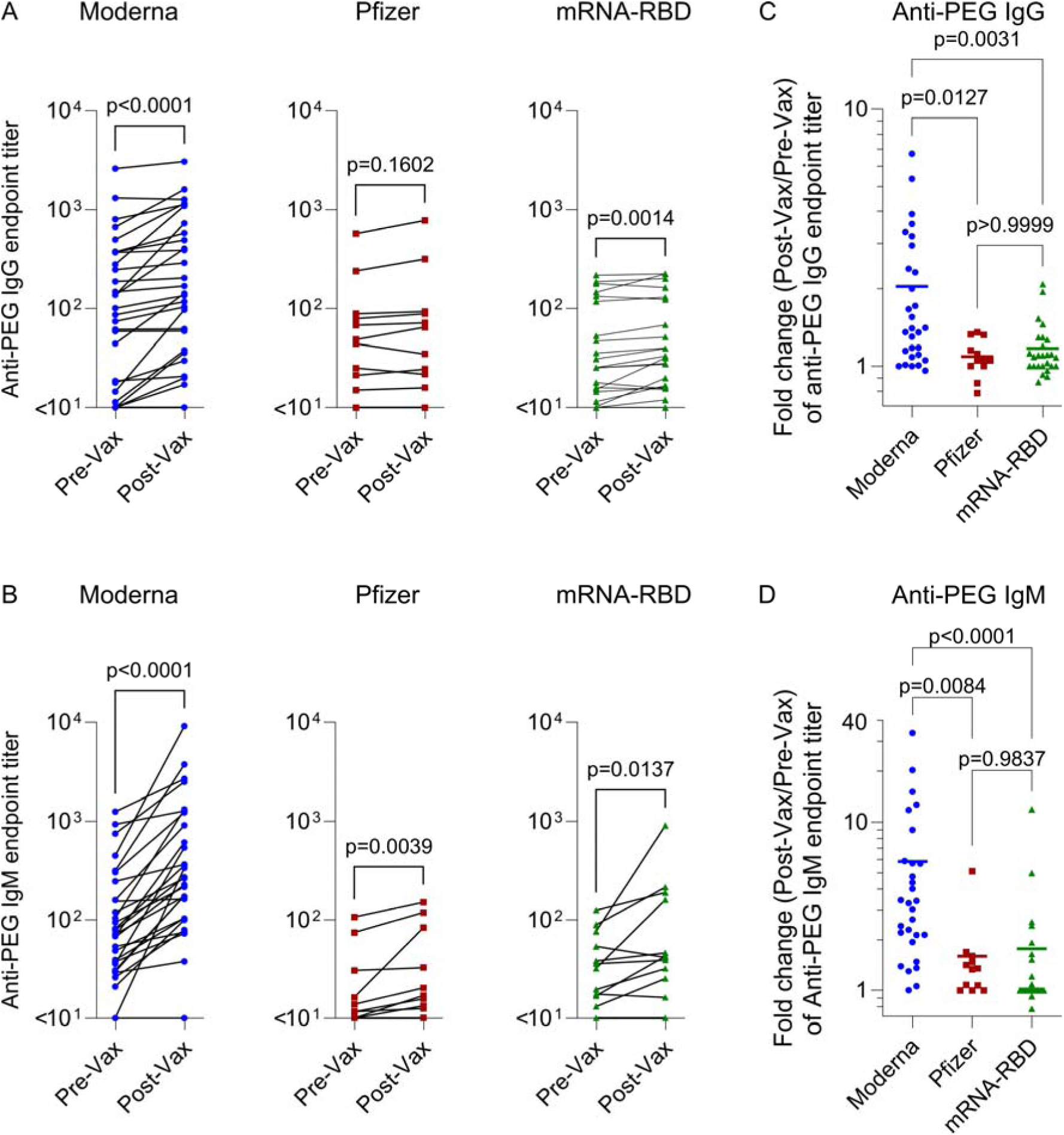
Comparison of anti-PEG antibody levels in human blood before and after Moderna, Pfizer, or mRNA-RBD vaccination. (A,B) Comparison of plasma anti-PEG IgG and IgM endpoint titers before vaccination (Pre-Vax) and after vaccination (Post-Vax) for the three vaccine types. (C,D) Cross-comparison of fold changes (Post-Vax/Pre-Vax) in anti-PEG IgG and IgM endpoint titers among the three vaccine types. In (C,D), each dot represents one participant, and the horizontal line indicates the mean. Statistical analysis was performed using the nonparametric Wilcoxon matched-pairs signed rank test in (A,B) and the nonparametric Kruskal–Wallis test with Dunn’s multiple comparisons in (C,D).

Vaccine immunogenicity was confirmed by significant increases in plasma anti-spike IgG titers after vaccination in all three cohorts (Fig. S6A–C). The magnitude of anti-spike IgG boosting did not differ across the three Moderna formulations (Fig. S6D), and anti-spike IgG fold changes were similar across the 10, 20, and 50 µg mRNA–RBD dose groups (Fig. S6E), indicating no significant formulation- or dose-dependent effects within the tested ranges. Notably, anti-spike antibodies may be less informative for the mRNA-RBD vaccine than for the Moderna and Pfizer vaccines, as the former encodes only the RBD, which is less than 20% of spike protein and presents a far more limited range of target antigens.

### Degradation patterns of Moderna vaccine mRNA

The prototype linkage RT-ddPCR assay above measures integrity across a single long span (37–42%) of spike mRNA, but it cannot determine whether integrity loss is biased toward particular regions of the transcript. To map degradation patterns along the spike transcript, we developed a ten-fragment linkage RT-ddPCR panel, in which each fragment is defined by two widely separated targets that are co-detected within the same droplet (Fig. 7A). The panel includes fragments shorter and longer than the original linkage assay (corresponding to fragment 6 in Fig. 7A) and covers both the 5′-proximal and 3′-proximal regions of the transcript. We applied this panel to serial plasma samples from six Moderna recipients (three BA.1 bivalent and three BA.5 bivalent), selected based on longitudinal sample availability.

**Figure 7.**
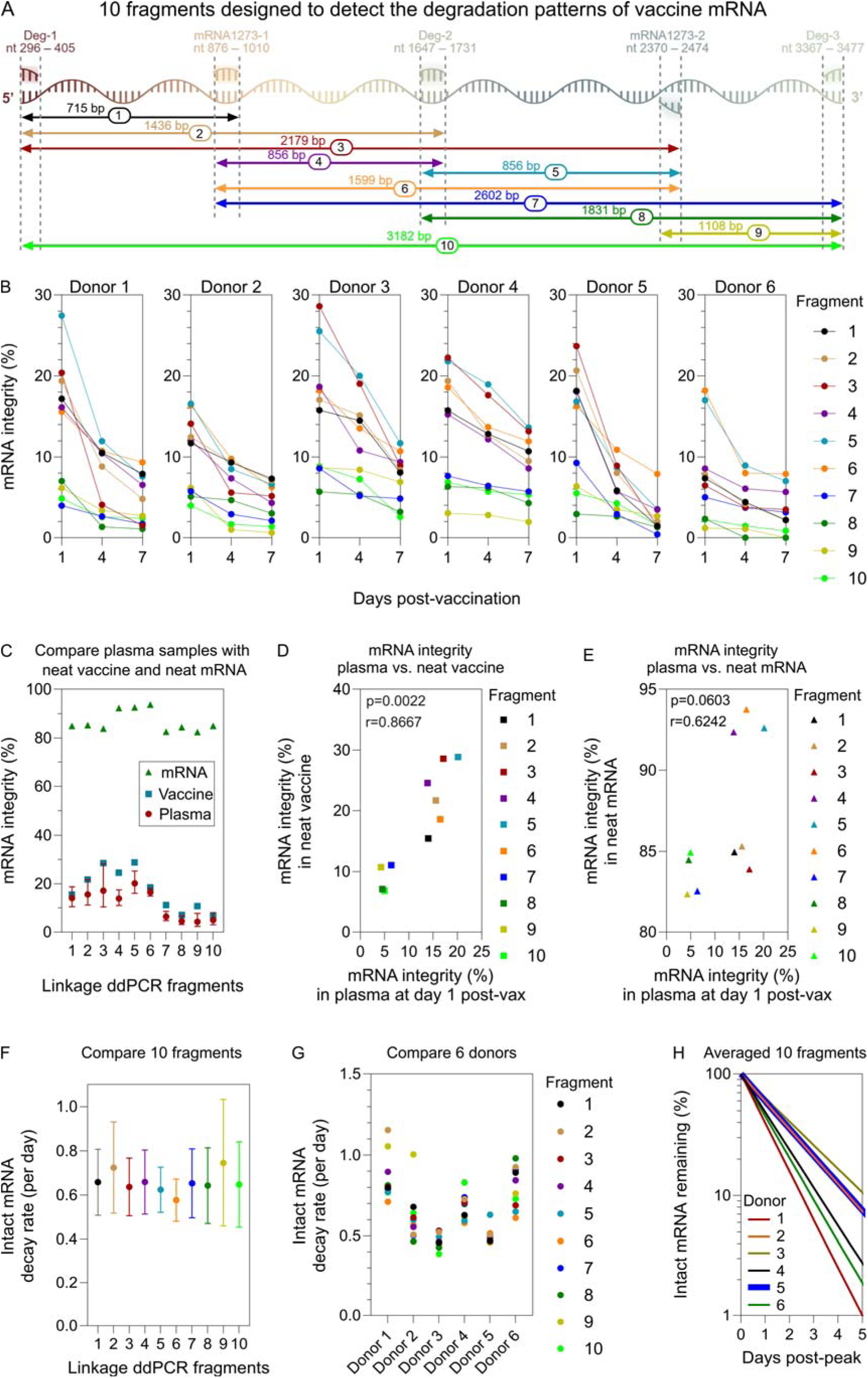
*In vivo* degradation patterns of Moderna vaccine mRNA in human blood evaluated using ten two-primer fragments in a duplex PCR assay. (A) Schematic illustration of the ten two-primer fragments, each targeting two regions of the Moderna vaccine mRNA sequence in the duplex ddPCR assay to assess degradation patterns of vaccine mRNA. (B) Vaccine mRNA integrity in plasma from six subjects at days 1, 4, and 7 post-Moderna vaccination (three subjects received the bivalent ancestral + BA.1 vaccine and three received the bivalent ancestral + BA.5 vaccine) assessed using the ten fragments. (C) Comparison of mRNA integrity across the ten fragments in plasma samples (day 1 post-vaccination), neat Moderna vaccine, and synthetic Moderna vaccine mRNA. (D,E) Spearman correlation analysis between mRNA integrity in plasma at day 1 post-vaccination and mRNA integrity in (D) neat Moderna vaccine or (E) synthetic Moderna vaccine mRNA. (F,G) Comparison of intact mRNA decay rates across (F) the ten fragments or (G) the six subjects. (H) Best-fit decay slopes of intact mRNA across six donors, with each data point representing the average decay rate calculated from ten individual fragments. For each donor, decay rates were estimated separately for each fragment, and the mean of these ten fragment-specific rates was used to represent the donor-level decay slope. As the decay slopes of donors 2 and 5 overlap, the curve of donor 5 was plotted with higher thickness than that of donor 2 to improve readability. In (C,F), mRNA integrity (%) and decay rates in plasma samples are shown as the mean with upper and lower bound of 95% confidence intervals calculated across six donors.

Intact (linked across the assayed span) Spike mRNA was detectable for all ten fragments in all six subjects over time (Fig. 7B). At day 1 post-vaccination, fragments nearest the 3’ end of the transcript (fragments 7–10) showed lower linkage (mean integrity 5.6%, range 1.2–9.3%) than mid-transcript fragments (fragments 4–6, mean integrity 17.6%, range 8.6–27.5%) or 5′-proximal fragments (fragments 1–3, mean integrity 16.6%, range 6.5–28.6%). Importantly, the day-1 integrity profile in plasma closely mirrored that measured in the administered vaccine product: the relative abundance of linked mRNA across fragments at day 1 (red circles in Fig. 7C) significantly correlated with fragment linkage in the original Moderna formulation (blue squares in Fig. 7C, Fig. 7D). As a reference, synthetic spike mRNA, synthesized based on the putative Moderna vaccine mRNA sequence, was largely intact across all ten fragments (green triangles in Fig. 7C, mean integrity 86.7%, range 82.4–93.7%) and did not show a significant correlation with the fragment-by-fragment profile observed in blood (Fig. 7E).Together, these data suggest that (i) the pattern of fragment integrity detected in blood at day 1 largely reflects the formulation’s starting integrity profile; (ii) there was minimal further degradation between vaccination and one day post-vaccination, when mRNA–LNPs travel *in vivo* into the blood; (iii) the integrity loss of vaccine product is biased toward the 3′-proximal regions of the transcript. Interestingly, the longest (near full-length) fragment (fragment 10) displayed comparatively low linkage at day 1 (mean integrity 5.4%, range 2.3–8.8%), a notable observation given that full-length transcripts are expected to contribute most directly to spike expression. Vaccine immunogenicity was nonetheless confirmed by significant boosting of anti-spike IgG after Moderna vaccination (Fig. S6A,D).

To assess whether mRNA degrades in a region-specific manner while circulating in the human blood *in vivo*, we tracked each fragment over the first week post-vaccination (Fig. 7B). All ten linked fragments declined over time, but the decay rates were similar across fragments (mean 0.65, range 0.58–0.72 dayL¹; Fig. 7F), suggesting no positional “hotspot” of transcript loss (or protection) *in vivo*. In contrast, decay rates clustered by individual (Fig. 7G): fragment slopes were more similar within a subject than between subjects. Across six subjects and ten fragments, ∼92% of the variance in decay rates was attributable to subject-to-subject differences, whereas ∼8% was attributable to fragment identity (based on mixed-effects modeling). We then plotted the mean decay rate across the 10 fragments for each individual against time (days post-peak), revealing substantial interpersonal differences (Fig. 7H).

## Discussion

The rational improvement of mRNA vaccines and therapeutics requires a precise understanding of how mRNA–LNP components behave *in vivo* in humans, particularly regarding their systemic distribution persistence, and transcript integrity over time. In this study, we performed a head-to-head comparison of three mRNA–LNP vaccine platforms: Moderna, Pfizer, and mRNA–RBD vaccine, across 73 participants and 327 serial blood samples. By quantifying total vaccine mRNA, long-range linked (“intact”) mRNA, platform-specific ionizable lipids, and antibody responses, we identified distinct platform-level differences. While peak circulating mRNA concentrations were broadly comparable between Moderna and Pfizer, their decay kinetics diverged significantly: Moderna displayed the fastest clearance, Pfizer an intermediate rate, and the mRNA–RBD the slowest. Notably, while ionizable lipid kinetics followed this same rank order, the total lipid exposure (AUC) was highest for Pfizer. Furthermore, our linkage RT-ddPCR analysis revealed that intact mRNA declined more rapidly following Moderna vaccination than Pfizer vaccination, highlighting platform-specific differences in the stability of long-range transcript integrity in circulation. Finally, while all platforms robustly boosted anti-spike IgG, Moderna induced significantly larger boosts in anti-PEG IgG and IgM than the Pfizer or mRNA–RBD vaccines.

These findings have important implications for the clinical application and evaluation of mRNA platforms. Conceptually, the detection of vaccine-derived mRNA and lipid in the blood likely represents a “spill-over” event from the injection site or draining lymph nodes. Assuming an average adult blood volume of 5 L, we estimate that a mean of 1.4% (range 0.01–5.3%) of the Moderna vaccine dose enters systemic circulation by day 1 post-vaccination. By day 6–7, only a minute fraction (mean 0.15%) remains. In contrast, higher proportions of the Pfizer and mRNA–RBD vaccines appear to remain in circulation at later time points (mean 1.9% and 0.5% at day 6–7, respectively,), likely driven by their slower decay rates relative to Moderna.

A critical mechanistic insight from this study is the platform-specific coupling of mRNA and lipid kinetics. We previously reported that intact Moderna mRNA and its ionizable lipid (SM-102) decay at almost identical rates, suggesting that the mRNA circulates largely protected within LNPs^7^. This study confirms that finding for Moderna. However, we observed a decoupling of these kinetics for the Pfizer vaccine, where the ionizable lipid (ALC-0315) persisted significantly longer than the intact mRNA. This suggests that Pfizer-derived lipids may circulate as “empty” or partially degraded LNPs after the mRNA cargo has been cleared. While the clinical impact of systemic vaccine distribution remains to be fully defined, circulating components presumably traffic to secondary lymphoid tissues, such as the spleen, potentially initiating immune responses or reactogenicity at distant sites. Although our study was not powered to robustly link circulating vaccine levels with side effects, these pharmacokinetic parameters offer valuable surrogate markers for rationally assessing vaccine safety and reactogenicity profiles in future formulations.

The divergence in “intact” mRNA stability, specifically the two-fold longer half-life of intact Pfizer mRNA compared to Moderna, is of particular interest. While our measurements are restricted to blood, it is plausible that similar stability profiles exist within lymph nodes. Theoretically, prolonged antigen availability could amplify humoral immunity by synchronizing antigen delivery with the developing germinal center response, a mechanism shown to enhance antibody titers in animal models^16^.

Furthermore, this study advances the characterization of mRNA degradation through a novel suite of linkage RT-ddPCR assays mapping the entire spike transcript. We identified a distinct positional bias in degradation: the 3’ end of the spike mRNA was most prone to loss, both in the vaccine formulation and in early post-vaccination blood samples. Remarkably, as little as 2.3% of the spike mRNA appeared intact across our largest fragment (spanning 3,182 bp, or ∼83% of the coding region) in early blood samples. This indicates that the vast majority of circulating mRNA exists as fragments. Strategies to stabilize the 3’ region and improve the proportion of full-length transcripts could represent a tangible target for enhancing the potency and efficiency of future mRNA vaccines.

Our data also shed light on the drivers of PEG immunogenicity. The observation that Moderna induced greater anti-PEG responses than Pfizer raises the question of whether this is driven by dose (50 µg vs. 30 µg) or LNP composition. Interestingly, we observed no dose-dependent increase in PEG immunogenicity for the mRNA–RBD vaccine across the 10, 20, and 50 µg cohorts, suggesting that dose alone may not be the primary determinant. Given that the mRNA–RBD and Moderna vaccines utilize the same PEG-lipid (PEG2000-DMG) yet elicit different responses, and that Moderna (SM-102) induced higher anti-PEG antibody titers than mRNA–RBD (Dlin-MC3-DMA), the specific ionizable lipid likely plays an important role. Ionizable lipids are known to possess adjuvant properties that promote antigen immunogenicity^17^. Our data suggest this adjuvant effect may extend to the induction of anti-PEG antibodies. Larger cohort and animal studies are warranted to confirm this hypothesis.

Broadly, typical vaccine evaluations prioritize immune endpoints and reactogenicity, often overlooking the physical pharmacokinetics of the vaccine components themselves. We advocate for a more balanced evaluation framework that includes the systemic distribution, persistence, and mRNA integrity of the vaccine. Our approach, initially driven by PCR-based mRNA detection, now expanded to LC–MS-based lipid detection and linkage ddPCR-enabled mRNA integrity mapping, offers a granular view of vaccine behavior *in vivo*. Recent work has detected PEG in human plasma samples^18^, suggesting it should be possible to measure the PEG-lipids of mRNA–LNP vaccines in plasma. Similar analyses could be attempted with other vaccine modalities.

We acknowledge several limitations in our study that should be explored in future studies. The sampling frequency for the investigational mRNA–RBD cohort was lower than for the licensed vaccines, reducing the temporal resolution of that comparison. Additionally, the heterogeneity in participants’ prior infection and vaccination history may introduce variability, though this reflects the real-world landscape of current vaccine administration. While this is the largest study of its kind, larger cohorts would allow for more robust correlation analyses between blood vaccine kinetics, immunogenicity, and reactogenicity.

Technically, while our linkage RT-ddPCR provides the most comprehensive map of transcript integrity to date, it does not offer single-nucleotide resolution; deep sequencing remains challenging due to the abundance of host RNA in blood. Finally, our analysis was restricted to blood; future studies examining biodistribution in draining lymph nodes, spleen, and mucosal sites, would provide a more complete anatomical picture.

In conclusion, we document and compare the *in vivo* kinetics of SARS-CoV-2 mRNA–LNP vaccine components from three vaccine platforms in humans. By establishing a quantitative framework to benchmark systemic persistence and transcript integrity, this work provides critical insights to guide the rational design of next-generation mRNA vaccines with improved stability, immunogenicity, and safety.

## Methods

### Ethics statement

The study protocols were approved by the University of Melbourne Human Research and Ethics Committee (approvals no. 2056689), and all associated procedures were carried out in accordance with the approved guidelines. The investigational RBD mRNA–LNP vaccine clinical study was approved by the Royal Melbourne Hospital Human Research Ethics Committee and was conducted under the Clinical Trial Notification (CTN) Scheme (CTN ID: 04968-1) administered by the Australian Therapeutic Goods Administration and registered at www.clinicaltrials.gov (NCT05272605). All participants provided written informed consent in accordance with the Declaration of Helsinki.

### Participants and sample collection

We recruited 29 participants who received a Moderna mRNA-1273 SPIKEVAX booster vaccination and 12 participants who received a Pfizer/BioNTech BNT162b2 vaccination. In addition, we analyzed plasma samples from 32 participants enrolled in a previously reported investigational RBD mRNA–LNP vaccine clinical trial^2^. Demographic characteristics and SARS-COV-2 vaccination history for all 73 participants are summarized in Table S1. Whole blood was collected into tubes containing sodium heparin or ethylenediaminetetraacetic acid (EDTA) anticoagulant, or into serum-separating tubes. Plasma and serum were aliquoted and stored at −80 °C.

### Quantification of COVID-19 vaccine mRNA

COVID-19 vaccine mRNA in plasma was quantified by reverse transcription droplet digital polymerase chain reaction (RT-ddPCR) as previously reported^7^. Total RNA was isolated from 140 µL of EDTA-anticoagulated plasma or serum using the QIAmp RNA Extraction Kit (Qiagen). RNA (10 µL) was reverse transcribed using Superscript III Reverse Transcriptase according to the manufacturers’ instructions (Invitrogen). Primers and probes were designed based on the putative vaccine mRNA sequences (Tables S3 and S4) and synthesized by Integrated DNA Technologies (USA). These primer/probe sets were specific for the codon-modified vaccine mRNA sequences and did not amplify wild-type SARS-CoV-2 spike (S)-gene.

ddPCR was performed on a QX200 system (Bio-Rad). Each 24-µL reaction contained 12 µL of 2× ddPCR Supermix for Probes (no dUTP, Bio-Rad), 5 µL of cDNA, and 5 µL of primer/probe mix. Droplets were generated using DG8 droplet generator cartridges (Bio-Rad) and a QX200 droplet generator (Bio-Rad). PCR was performed on a C1000 Touch thermal cycler (Bio-Rad) with the following program: 95°C for 10 min; 40 cycles of 94°C for 30 s and 60°C for 60 s; and 98°C for 10 min, with a ramp rate of 2°C/s. Droplets were read on a QX200 droplet reader (Bio-Rad), and data were analyzed using QuantaSoft software v1.7.4.

Vaccine mRNA from the original vaccine product was used as a positive control and to set the positive droplet threshold and to generate linear standard curves. Vaccine was spiked into negative human plasma (collected pre-vaccination), followed by RNA extraction and reverse transcription as described above. cDNA was serially diluted 10-fold in nuclease-free water and analyzed by ddPCR. Copy numbers of the vaccine mRNA in the ddPCR reaction were used to derive the copy number per µL of plasma. The linear PCR standard curve (Fig. S1D–G) was derived in GraphPad Prism 10 with relative weighting (weighting by 1/*y*^2^) and used to covert mRNA concentration from copies µL^−1^ to ng mL^−1^.

### Quantification of ionizable lipids

Ionizable lipids (SM-102, ALC-0315, and Dlin-MC3-DMA) were quantified by targeted liquid chromatography–mass spectrometry (LC–MS) as previously reported^7^. For sample preparation, 50 μL of diluted plasma (90% plasma, prepared by mixing 45 μL neat plasma with 5 μL phosphate-buffered saline (PBS)) was extracted with 500 μL of 1-butanol/methanol (1:1, v/v). Then, samples were vortexed for 10 s and sonicated for 60 min in a water bath at 20 °C. To generate linear standard curves, the original vaccine product (with known ionizable lipid concentration) was serially diluted in PBS, and 5 µl of diluted vaccine was spiked into 45 μL of negative human plasma (collected pre-vaccination), followed by the same extraction procedure as described above. Samples were subsequently centrifuged at 16,000 × g for 20 min, and 100 μL of the resulting supernatant was transferred to glass LC autosampler vials (Phenomenex, USA) for analysis.

Samples were analyzed using a Shimadzu 8050 triple quadrupole mass spectrometer coupled to a Shimadzu Nexera X2 liquid chromatography system. Sample components were separated on an Agilent RRHD Eclipse Plus C18 column (2.1 × 1000 mm, 1.8 μm; Agilent Technologies, USA) over a 15 min gradient using mobile phase A (6:4 water/acetonitrile containing 10 mM ammonium acetate and 5 μM medronic acid) and mobile phase B (9:1 isopropanol/acetonitrile containing 10 mM ammonium acetate). The solvent gradient was programmed as follows [time (min), B (%)]: [0, 5], [2, 5], [10, 98], [12, 98], [12.5, 5], [15, 5]. The solvent flow rate was maintained at 0.3 mL min^−1^, and the eluent was diverted to waste for the first 5 min. The autosampler chamber and column oven were maintained at 10 and 40 °C, respectively.

Analytes were ionized by electrospray ionization in positive ion mode and detected using multiple reaction monitoring (MRM) strategy (Table S8). Nebulizing gas flow rate was 2 L min^−1^, and heating and drying gas flow rates were 10 L min^−1^. The interface temperature and voltage were maintained at 375 °C and 4 kV, respectively. Collision energies and MRM parameters were optimized using the in-built Shimadzu LabSolutions MRM optimization tool.

Raw data were processed using Skyline (v23.1). Analytes were quantified by integrating peak areas from the summed MRM transitions. Standard curves (Fig. S3D–G) were fitted using weighted linear regression (1/*x*^2^) and used to calculate the ionizable lipid concentrations.

### Linkage RT-ddPCR to measure relative levels of intact mRNA

We performed linkage RT-ddPCR as originally developed by Hanna et al. to quantify vaccine mRNA integrity in human breast milk^19^ and as subsequently adapted by us to quantify vaccine mRNA integrity in human plasma^7^. Linkage RT-ddPCR uses a duplex ddPCR design in which two widely separated regions of the same spike mRNA molecule are detected simultaneously using probes labeled with different fluorophores (FAM and HEX). Droplets that are double-positive (FAM+HEX) indicate that both target regions were present within the same partition and therefore remain physically linked on the same RNA molecule across the assayed span (i.e., “intact” across that region). In contrast, droplets that are single-positive (FAM-only or HEX-only) are consistent with fragmented mRNA species that contain only one of the two target regions.

Linkage duplex ddPCR reactions were prepared and run using the same droplet generation, thermal cycling, droplet reading, and analysis workflow described above for total vaccine mRNA quantification, except that the linkage primer/probe sets (Tables S5–S6) were combined in a single duplex reaction (FAM and HEX). Data were analyzed in QuantaSoft v1.7.4. QuantaSoft was used to compute the linkage number (linked copies), defined as the excess number of double-positive droplets above that expected from random co-occupancy of unlinked targets (i.e., correcting for chance colocalization).

Percent linkage (the “intact” fraction across the assayed span) was calculated as:

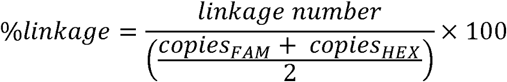

where *copies_FAM_* and *copies_HEX_* are the single-target concentrations reported for each channel in QuantaSoft.

To map integrity patterns along the spike transcript, we expanded the original linkage assay into a ten-fragment linkage RT-ddPCR panel, using distinct duplex primer/probe pairs to quantify linkage across ten different transcript spans (Fig. 7A; Table S7). Each fragment was analyzed using the same duplex linkage workflow described above. As a reference control, synthetic spike mRNA was produced by Messenger Bio (Australia) based on the putative Moderna vaccine mRNA sequence and assayed alongside vaccine and plasma samples.

### Quantification of anti-PEG antibody

Anti-PEG IgG and IgM were quantified by enzyme-linked immunosorbent assay (ELISA) as previously described.^14^ Briefly, eight-arm PEG-NHL (40 kDa; 200 µg/mL; JenKem Technology, USA) in PBS was coated onto MaxiSorp 96-well plates (Nunc, Denmark) for 18 h at 4°C. Plates were washed four times with PBS and blocked with 5% (w/v) skim milk in PBS for 22 h. Serially diluted human plasma samples (prepared in 5% skim milk in PBS) were added in duplicate and incubated for 1 h at 22°C. Plates were washed twice with 0.1% 3-[(3-cholamidopropyl)-dimethylammonio]-1-propanesulfonate (CHAPS; Sigma-Aldrich, USA) in PBS and then four times with PBS before adding horseradish peroxidase (HRP)-conjugated anti-human IgG (Dako Agilent, USA; 1:20,000 dilution) or HRP-conjugated anti-human IgM (Jackson ImmunoResearch Laboratories, USA; 1:10,000 dilution) for 1 h at 22°C. Plates were washed as above and developed with 3,3′,5,5′-tetramethylbenzidine (TMB) liquid substrate (Sigma-Aldrich, USA). Reactions were stopped with 0.16 M HLSOL, and absorbance was read at 450 nm. Endpoint titers were defined as the reciprocal plasma dilution yielding a signal two-fold above background, calculated in GraphPad Prism 10 using a fitted curve (4-parameter log regression) and reported as the mean of duplicates. Background signals were determined by applying plasma (1:10 dilution in 5% skim milk) to non-PEG-coated wells and processing in parallel.

### Quantification of anti-spike IgG

Plasma IgG binding to the ancestral SARS-CoV-2 spike protein was measured by ELISA following an established protocol^7^. Briefly, 96-well Maxisorp plates were coated overnight at 4 °C with recombinant HexaPro^20^ spike protein (2 μg mL^−1^). Plates were washed twice with PBS containing 0.05% Tween-20 and once with PBS, then blocked with PBS containing 1% fetal calf serum (FCS) for 1 h at 22°C. Plasma samples were serially diluted 4-fold (1:100 to 1:1,638,400), added in duplicate, and incubated for 2 h at 22°C. Following incubation, plates were washed four times with PBS containing 0.05% Tween-20 and then twice with PBS. Bound IgG was detected using HRP-conjugated anti-human IgG antibody (Agilent, USA; 1:20,000 dilution). Plates were developed with TMB substrate, the reaction was stopped with 0.16 M HLSOL, and absorbance was read at 450 nm. Endpoint titers were calculated using GraphPad Prism 10 as the reciprocal serum dilution yielding a signal two-fold above background using a fitted curve (4 parameter log regression) and reported as the mean of duplicates. Background signals were determined by averaging the signals from all plasma samples at a dilution of 1:1,638,400.

### Estimating the decay rates

Decay rates of mRNA and lipid species in plasma were estimated using a linear mixed-effects model with *days post-peak* and *vaccine type* (Moderna, Pfizer, mRNA–RBD) as fixed effects. Subject-specific random intercepts and slopes were included to account for between-individual variability in baseline levels and decay rates. To represent exponential decay, response variables were log-transformed prior to analysis. Observations below the lower limit of quantitation (LLOQ) were treated as left censored. Random effects were assumed to follow a bivariate normal distribution, and residual errors were modeled as independent and normally distributed. Model fitting was performed using maximum likelihood estimation implemented in the *lmec* package in *R* (version 4.4.0). Differences in decay rates among response types were assessed using likelihood ratio tests.

### Statistical analysis

Decay rates of total mRNA (Fig. 2E), ionizable lipid (Fig. 3E), and intact mRNA (Fig. 4E) were compared using likelihood ratio tests in R (v4.4.0). Comparisons across the three vaccine platforms for mRNA concentration at day 6–7 (Fig. 2D), post-peak mRNA AUC (Fig. 2F), ionizable lipid concentration at day 6–7 (Fig. 3D), ionizable lipid AUC (Fig. 3F), and fold changes in anti-PEG IgG and IgM titers (Fig. 6C,D) were performed using nonparametric Kruskal–Wallis test with Dunn’s multiple comparisons in GraphPad Prism 10. Intact mRNA AUC was compared between Moderna and Pfizer (Fig. 4F) using the nonparametric Mann−Whitney *U* test in GraphPad Prism 10. Paired comparison of anti-PEG IgG and IgM titers pre- versus post-vaccination (Fig. 6A,B) and anti-spike IgG titers pre-versus post-vaccination (Fig S6A–C) were performed using the nonparametric Wilcoxon matched-pairs signed rank test in GraphPad Prism 10. Associations between mRNA integrity in plasma at day 1 post-vaccination and mRNA integrity in neat Moderna vaccine or synthetic Moderna vaccine mRNA (Fig. 7D,E) were assessed using nonparametric Spearman correlation in GraphPad Prism 10. Comparisons of AUCs for total mRNA, ionizable lipid, intact mRNA across Moderna formulations and across mRNA-RBD dose groups (Fig. S2A–E), as well as fold changes in anti-PEG and anti-spike antibody titers across Moderna formulations or mRNA–RBD dose groups (Fig. S5A–D and Fig. S6D,E) were performed using the Kruskal–Wallis test with Dunn’s multiple comparisons in GraphPad Prism 10.

## Supporting information

Supporting Informaion

## Data Availability

All data produced in the present study are available upon reasonable request to the authors

## Author Contributions

We annotate author contributions using the CRediT Taxonomy labels. Conceptualization—Y.J. and S.J.K; Investigation—Y.J., S.L., and T.H.A.; Methodology—S.J.K., Y.J., T.H.A., S.L., and M.G.L.; Formal analysis—A.R. and M.P.D.; Funding acquisition—S.J.K., Y.J., and M.P.D.; Resources—S.J.K., Y.J., M.P.D., J.A.J., A.K.W., G.D., D.I.G., T.N., and C.W.P.; Supervision—S.J.K and Y.J.; Writing–original draft—S.J.K and Y.J.; Writing–review & editing—all co-authors.

## Acknowledgment

We thank the participations for the generous involvement and provision of samples. We acknowledge the Melbourne Mass Spectrometry and Proteomics Facility for provision of liquid chromatography–mass spectrometry services. This study was supported by an Australian Research Council (ARC) Discovery Early Career Researcher Award grant (DE230101542 to Y.J.), the National Health and Medical Research Council (NHMRC) Investigator grants (GNT2016491 to S.J.K.; J.A.J.; A.K.W.; M.P.D.; GNT2008913 to D.I.G; GNT2043120 to Y.J.), Medical Research Future Fund (MRFF) Awards (2005846, 2005990) (D.I.G. and T.N.), and the Victorian Critical Vaccinees Collection (VC^2^) COVID-19 Research Seed Funding Grant (Y.J.). Fig. 1 and Fig. 7A were created with BioRender.com.

## Notes

### Competing Interest Statement

The authors have declared no competing interest.

### Clinical Trial

NCT05272605

### Author Declarations

The study protocols were approved by the University of Melbourne Human Research and Ethics Committee (approvals no. 2056689), and all associated procedures were carried out in accordance with the approved guidelines. The investigational RBD mRNA-LNP vaccine clinical study was approved by the Royal Melbourne Hospital Human Research Ethics Committee and was conducted under the Clinical Trial Notification (CTN) Scheme (CTN ID: 04968-1) administered by the Australian Therapeutic Goods Administration and registered at www.clinicaltrials.gov (NCT05272605). All participants provided written informed consent in accordance with the Declaration of Helsinki.

