## Supporting Informaion for "In Vivo Blood Kinetics and Transcript Integrity of Three mRNA–Lipid Nanoparticle Vaccines in Humans"

**Table S1.** Demographic information and SARS-COV-2 immunization history of 73 participants who received either Moderna mRNA-1273 (Spikevax), Pfizer/BioNTech BNT162b2 (Comirnaty), or mRNA–RBD SARS-CoV-2 mRNA vaccination.

| Participant ID | Age | Sex | SARS-COV-2 Vaccine history | Moderna Spikevax vaccination | | |
|---|---|---|---|---|---|---|
| | | | | Type | mRNA dose (µg) | Days since last COVID-19 vaccination |
| 1 | 36−40 | M | Pfizer (×3) | Ancestral & BA.1 | 50 | 401 |
| 2 | 26−30 | F | Pfizer (×3) | Ancestral & BA.1 | 50 | 343 |
| 3 | 31−35 | F | Pfizer (×3) | Ancestral & BA.1 | 50 | 424 |
| 4 | 36−40 | M | AstraZeneca (×2) + Moderna (×1) + Pfizer (×1) | Ancestral & BA.1 | 50 | 191 |
| 5 | 61−65 | M | AstraZeneca (×2) + Moderna (×1) + Novavax (×1) | Ancestral & BA.1 | 50 | 192 |
| 6 | 26−30 | F | Pfizer (×3) | Ancestral & BA.1 | 50 | 346 |
| 7 | 56−60 | F | AstraZeneca (×2) + Moderna (×1) | Ancestral & BA.5 | 50 | 412 |
| 8 | 61−65 | F | AstraZeneca (×2) + Pfizer (×2) | Ancestral & BA.5 | 50 | 256 |
| 9 | 56−60 | F | AstraZeneca (×2) + Moderna (×1) | Ancestral & BA.5 | 50 | 486 |
| 10 | 21−25 | F | AstraZeneca (×2) + Moderna (×2) | Ancestral & BA.5 | 50 | 139 |
| 11 | 61−65 | M | Novavax (×1) + Pfizer (×3) | Ancestral & BA.5 | 50 | 270 |
| 12 | 31−35 | F | Pfizer (×2) + Moderna (×1) | Ancestral & BA.5 | 50 | 432 |
| 13 | 31−35 | M | Pfizer (×3) | Ancestral & BA.5 | 50 | 433 |
| 14 | 26−30 | F | Pfizer (×3) | Ancestral & BA.5 | 50 | 433 |
| 15 | 21−25 | F | AstraZeneca (×3) | Ancestral & BA.5 | 50 | 419 |
| 16 | 26−30 | M | Pfizer (×3) | Ancestral & BA.5 | 50 | 457 |
| 17 | 31−35 | F | Pfizer (×1) + Moderna (×2) | Ancestral & BA.5 | 50 | 224 |

| | | | | | | |
|---|---|---|---|---|---|---|
| 18 | 61−65 | F | AstraZeneca (×2) + Moderna (×1) | Ancestral & BA.5 | 50 | 496 |
| 19 | 66−70 | M | AstraZeneca (×2) + Moderna (×1) | Ancestral & BA.5 | 50 | 380 |
| 20 | 61−65 | M | Pfizer (×3) + Moderna (×1) | XBB.1.5 | 50 | 355 |
| 21 | 31−35 | F | Moderna (×3) + Pfizer (×1) | XBB.1.5 | 50 | 431 |
| 22 | 26−30 | F | Pfizer (×3) + Moderna (×1) | XBB.1.5 | 50 | 508 |
| 23 | 26−30 | F | Pfizer (×3) + Moderna (×1) | XBB.1.5 | 50 | 460 |
| 24 | 36−40 | M | Pfizer (×3) + Moderna (×1) | XBB.1.5 | 50 | 460 |
| 25 | 31−35 | F | Moderna (×2) + Pfizer (×1) | XBB.1.5 | 50 | 832 |
| 26 | 56−60 | F | Pfizer (×3) | XBB.1.5 | 50 | 872 |
| 27 | 21−25 | F | Covishield (×3) + Moderna (×1) | XBB.1.5 | 50 | 362 |
| 28 | 26−30 | F | Sinopharm (×3) | XBB.1.5 | 50 | 740 |
| 29 | 26−30 | F | AstraZeneca (×2) + Moderna (×2) | XBB.1.5 | 50 | 487 |
| Summary | Median 33 (range 24−70) | F 69% | Median 3 vaccines (range 3−4) | BA.1 (n=6); BA.5 (n=13); XBB.1.5 (n=10) | 50 µg (n=29) | Median 424 (range 139−872) |

| **Participant ID** | **Age** | **Sex** | **SARS-COV-2 Vaccine history** | **Pfizer/BioNTech Comirnaty vaccination** | | |
|---|---|---|---|---|---|---|
| | | | | **Type** | **mRNA dose (µg)** | **Days since last COVID-19 vaccination** |
| 30 | 51−55 | F | None | Ancestral | 30 | N.A. |
| 31 | 61−65 | M | None | Ancestral | 30 | N.A. |
| 32 | 21−25 | F | None | Ancestral | 30 | N.A. |
| 33 | 51−55 | M | None | Ancestral | 30 | N.A. |
| 34 | 66−70 | F | None | Ancestral | 30 | N.A. |
| 35 | 56−60 | F | None | Ancestral | 30 | N.A. |
| 36 | 56−60 | M | None | Ancestral | 30 | N.A. |
| 37 | 56−60 | F | None | Ancestral | 30 | N.A. |
| 38 | 21−25 | F | None | Ancestral | 30 | N.A. |
| 39 | 51−55 | M | None | Ancestral | 30 | N.A. |
| 40 | 31−35 | F | None | Ancestral | 30 | N.A. |
| 41 | 46−50 | F | None | Ancestral | 30 | N.A. |
| Summary | Median 55 (range 23−67) | F 67% | None | Ancestral (n=12) | 30 µg (n=12) | N.A. |

| Participant ID | Age | Sex | SARS-COV-2 Vaccine history | mRNA–RBD vaccination | | |
|---|---|---|---|---|---|---|
| | | | | Type | mRNA dose (μg) | Days since last COVID-19 vaccination |
| 42 | 61−65 | M | AstraZeneca (×2) + Pfizer (×1) | B.1.351 | 10 | 111 |
| 43 | 46−50 | F | Pfizer (×3) | B.1.351 | 10 | 99 |
| 44 | 46−50 | F | Pfizer (×3) | B.1.351 | 10 | 165 |
| 45 | 31−35 | F | AstraZeneca (×2) + Pfizer (×1) | B.1.351 | 10 | 148 |
| 46 | 61−65 | F | AstraZeneca (×2) + Pfizer (×1) | B.1.351 | 10 | 169 |
| 47 | 56−60 | M | AstraZeneca (×2) + Pfizer (×1) | B.1.351 | 10 | 151 |
| 48 | 51−55 | F | AstraZeneca (×2) + Moderna (×1) | B.1.351 | 10 | 161 |
| 49 | 56−60 | M | AstraZeneca (×2) + Pfizer (×1) | B.1.351 | 10 | 180 |
| 50 | 46−50 | M | AstraZeneca (×2) + Moderna (×1) | B.1.351 | 10 | 194 |
| 51 | 46−50 | F | Pfizer (×3) | B.1.351 | 10 | 192 |
| 52 | 31−35 | M | Pfizer (×3) | B.1.351 | 10 | 196 |
| 53 | 61−65 | F | AstraZeneca (×2) + Pfizer (×1) | B.1.351 | 10 | 188 |
| 54 | 31−35 | M | Pfizer (×3) | B.1.351 | 10 | 156 |
| 55 | 36−40 | F | Pfizer (×3) | B.1.351 | 10 | 187 |
| 56 | 31−35 | F | Pfizer (×3) | B.1.351 | 10 | 248 |
| 57 | 46−50 | M | Pfizer (×3) | B.1.351 | 10 | 213 |
| 58 | 56−60 | M | AstraZeneca (×2) + Moderna (×1) | B.1.351 | 20 | 216 |
| 59 | 41−45 | F | Pfizer (×3) | B.1.351 | 20 | 277 |
| 60 | 16−20 | F | Pfizer (×3) | B.1.351 | 20 | 192 |
| 61 | 26−30 | F | Pfizer (×3) | B.1.351 | 20 | 209 |
| 62 | 26−30 | F | Pfizer (×3) | B.1.351 | 20 | 232 |
| 63 | 26−30 | F | Pfizer (×3) | B.1.351 | 20 | 224 |
| 64 | 46−50 | F | Pfizer (×3) | B.1.351 | 20 | 301 |
| 65 | 36−40 | M | AstraZeneca (×2) + Pfizer (×1) | B.1.351 | 20 | 222 |
| 66 | 26−30 | F | AstraZeneca (×2) + Moderna (×1) | B.1.351 | 50 | 241 |

| | | | | | | |
|---|---|---|---|---|---|---|
| 67 | 36−40 | M | Pfizer (×2) + Moderna (×1) | B.1.351 | 50 | 243 |
| 68 | 41−45 | M | Pfizer (×2) + Moderna (×1) | B.1.351 | 50 | 278 |
| 69 | 21−25 | M | AstraZeneca (×2) + Pfizer (×1) | B.1.351 | 50 | 270 |
| 70 | 41−45 | M | Pfizer (×3) | B.1.351 | 50 | 322 |
| 71 | 31−35 | F | Pfizer (×2) + Moderna (×1) | B.1.351 | 50 | 267 |
| 72 | 61−65 | F | AstraZeneca (×2) + Moderna (×1) | B.1.351 | 50 | 302 |
| 73 | 51−55 | F | AstraZeneca (×2) + Pfizer (×1) | B.1.351 | 50 | 288 |
| Summary | Median 45 (range 20−63) | F 59% | 3 vaccines (n=32) | B.1.351 (n=32) | 10 µg (n=16); 20 µg (n=8); 50 µg (n=8) | Median 211 (range 99−322) |

**Table S2** Four lipid components in the LNP formulation of Moderna mRNA-1273 (Spikevax), Pfizer/BioNTech BNT162b2 (Comirnaty), and the mRNA–RBD SARS-CoV-2 mRNA vaccines.

| Vaccine | Ionizable lipid | PEG-lipid | Phospholipid | Other lipids |
|---|---|---|---|---|
| Moderna (Spikevax) | SM-102 | PEG2000-DMG | DSPC | Cholesterol |
| Pfizer/BioNTech (Comirnaty) | ALC-0315 | ALC-0159 | DSPC | Cholesterol |
| mRNA–RBD | Dlin-MC3-DMA | PEG2000-DMG | DSPC | Cholesterol |

**Table S3.** Sequences of primer and probe designed specifically for the codon-modified vaccine mRNA sequences used in ddPCR assays to quantify Moderna mRNA-1273 (Spikevax) and Pfizer/BioNTech BNT162b2 (Comirnaty) vaccine mRNA levels in plasma (data shown in Figure 2A,B).

| Primer or probe | Sequence | Target region |
|---|---|---|
| AMmRNA-F | 5-'GAGCCTGCTGATCGTGAATAA-3' | nt 402 – 514 (113 bp) |
| AMmRNA-P | 5'-/56-FAM/TCAAGGTGT/ZEN/GCGAGTTCCAGTTCT/3IABkFQ/-3' | |
| AMmRNA-R | 5'-TCCAGCTCTTGTTGTTCTTGT-3' | |

**Table S4.** Sequences of primer and probe designed specifically for the codon-modified vaccine mRNA sequences used in ddPCR assays to quantify mRNA–RBD vaccine mRNA levels in plasma (data shown in Figure 2C).

| Primer or probe | Sequence | Target region |
|---|---|---|
| RBD-F | 5'-GATCGCCGACTACAACTACAA-3' | nt 404 – 537 (134 bp) |
| RBD-P | 5'-/56-FAM/AACAGCAAC/ZEN/AACCTGGACAGCAAG/3IBkFQ/-3' | |
| RBD-R | 5'-TCAGGTTGCTCTTCCTGAAC-3' | |

**Table S5.** Sequences of primer and probe designed specific to respective codon-modified vaccine mRNA sequence for duplex ddPCR to quantify the integrity of Moderna mRNA-1273 (Spikevax) vaccine mRNA in plasma (data shown in Figure 4A)

| Primer or probe | Sequence | Target region | mRNA integrity region |
|---|---|---|---|
| mRNA1273-1-F | 5'-GACCTTCCTGCTGAAGTACAA-3' | nt 876 – 1010 | nt 876 – 2474 (1599 bp) |
| mRNA1273-1-P | 5'-/56-FAM/ACCAAGTGC/ZEN/ACCCTGAAGAGCTTC/3IABkFQ/-3' | | |
| mRNA1273-1-R | 5'-AAGTTGCTGGTCTGGTAGATG-3' | | |
| mRNA1273-2-F | 5'-GTGGAGCAGGACAAGAACA-3' | nt 2370 – 2474 | |
| mRNA1273-2-P | 5'-/56-HEX/CGCCCAGGT/ZEN/GAAGCAGATCTACAA/3IBkFQ/-3' | | |
| mRNA1273-2-R | 5'-AGGATCTGGCTGAAGTTGAAG-3' | | |

**Table S6.** Sequences of primer and probe designed specific to respective codon-modified vaccine mRNA sequence for duplex ddPCR to quantify the integrity of Pfizer/BioNTech BNT162b2 (Comirnaty) vaccine mRNA in plasma (data shown in Figure 4B)

| Primer or probe | Sequence | Target Region | mRNA integrity region |
|---|---|---|---|
| BNT-1-F | 5'-CAGCTACCAGACACAGACAAA-3' | nt 2070 – 2192 | nt 2070 – 3473 (1404 bp) |
| BNT-1-P | 5'-/56-FAM/AGCCAGAGC/ZEN/ATCATTGCCTACACA/3IBkFQ/-3' | | |
| BNT-1-R | 5'-GCGATAGAGTTGTTGGAGTAGG-3' | | |
| BNT-2-F | 5'-GTGACACAGCGGAACTTCTA-3' | nt 3364 – 3473 | |
| BNT-2-P | 5'-/56-HEX//TCATCACCA/ZEN/CCGACAACACCTTCG/3IBkFQ/-3' | | |
| BNT-2-R | 5'-GGGTCGTACACGGTATTGTT-3' | | |

**Table S7.** Ten two-primer fragments used in duplex PCR assay to detect the degradation patterns of Moderna mRNA-1273 (Spikevax) vaccine mRNA in plasma (data shown in Figure 7A,B)

| Fragment | Primer or probe | Sequence | Target region | mRNA integrity region |
|---|---|---|---|---|
| 1 | Deg-1-F | 5-'CATCATCAGAGGCTGGATCTTC-3' | nt 296 – 405 | nt 296 – 1010 (715 bp) |
| | Deg-1-P | 5'-/56-FAM/CACACTGGA/ZEN/CAGCAAGACCCAGAG/3IABkFQ/-3' | | |
| | Deg-1-R | 5-'GAACTGGAACTCGCACACTT-3' | | |
| | mRNA1273-1-F | 5'-GACCTTCCTGCTGAAGTACAA-3' | nt 876 – 1010 | |
| | mRNA1273-1-P | 5'-/56-HEX/ACCAAGTGC/ZEN/ACCCTGAAGAGCTTC/3IABkFQ/-3' | | |
| | mRNA1273-1-R | 5'-AAGTTGCTGGTCTGGTAGATG-3' | | |
| 2 | Deg-1-F | 5-'CATCATCAGAGGCTGGATCTTC-3' | nt 296 – 405 | nt 296 – 1731 (1436 bp) |
| | Deg-1-P | 5'-/56-FAM/CACACTGGA/ZEN/CAGCAAGACCCAGAG/3IABkFQ/-3' | | |
| | Deg-1-R | 5-'GAACTGGAACTCGCACACTT-3' | | |
| | Deg-2-F | 5-'CACCAACCTGGTGAAGAACA-3' | nt 1647 – 1731 | |
| | Deg-2-P | 5'-/56-HEX/CGTGCTGAC/ZEN/CGAGAGCAACAAGAA/3IABkFQ/-3' | | |
| | Deg-2-R | 5-'CTGCTGAAAGGGCAGGAAT-3' | | |
| 3 | Deg-1-F | 5-'CATCATCAGAGGCTGGATCTTC-3' | nt 296 – 405 | nt 296 – 2474 (2179 bp) |
| | Deg-1-P | 5'-/56-FAM/CACACTGGA/ZEN/CAGCAAGACCCAGAG/3IABkFQ/-3' | | |
| | Deg-1-R | 5-'GAACTGGAACTCGCACACTT-3' | | |

| | | | | |
|---|---|---|---|---|
| | mRNA1273-2-F | 5'-GTGGAGCAGGACAAGAACA-3' | nt 2370 – 2474 | |
| | mRNA1273-2-P | 5'-/56-HEX/CGCCCAGGT/ZEN/GAAGCAGATCTACAA/3IBkFQ/-3' | | |
| | mRNA1273-2-R | 5'-AGGATCTGGCTGAAGTTGAAG-3' | | |
| 4 | mRNA1273-1-F | 5'-GACCTTCCTGCTGAAGTACAA-3' | nt 876 – 1010 | nt 876 – 1731 (856 bp) |
| | mRNA1273-1-P | 5'-/56-HEX/ACCAAGTGC/ZEN/ACCCTGAAGAGCTTC/3IABkFQ/-3' | | |
| | mRNA1273-1-R | 5'-AAGTTGCTGGTCTGGTAGATG-3' | | |
| | Deg-2-F | 5-'CACCAACCTGGTGAAGAACA-3' | nt 1647 – 1731 | |
| | Deg-2-P | 5'-/56-FAM/CGTGCTGAC/ZEN/CGAGAGCAACAAGAA/3IABkFQ/-3' | | |
| | Deg-2-R | 5-'CTGCTGAAAGGGCAGGAAT-3' | | |
| 5 | Deg-2-F | 5-'CACCAACCTGGTGAAGAACA-3' | nt 1647 – 1731 | nt 1647 – 2474 (828 bp) |
| | Deg-2-P | 5'-/56-FAM/CGTGCTGAC/ZEN/CGAGAGCAACAAGAA/3IABkFQ/-3' | | |
| | Deg-2-R | 5-'CTGCTGAAAGGGCAGGAAT-3' | | |
| | mRNA1273-2-F | 5'-GTGGAGCAGGACAAGAACA-3' | nt 2370 – 2474 | |
| | mRNA1273-2-P | 5'-/56-HEX/CGCCCAGGT/ZEN/GAAGCAGATCTACAA/3IBkFQ/-3' | | |
| | mRNA1273-2-R | 5'-AGGATCTGGCTGAAGTTGAAG-3' | | |
| 6 | mRNA1273-1-F | 5'-GACCTTCCTGCTGAAGTACAA-3' | nt 876 – 1010 | nt 876 – 2474 (1599 bp) |
| | mRNA1273-1-P | 5'-/56-FAM/ACCAAGTGC/ZEN/ACCCTGAAGAGCTTC/3IABkFQ/-3' | | |

| | | | | |
|---|---|---|---|---|
| | mRNA1273-1-R | 5'-AAGTTGCTGGTCTGGTAGATG-3' | | |
| | mRNA1273-2-F | 5'-GTGGAGCAGGACAAGAACA-3' | | |
| | mRNA1273-2-P | 5'-/56-HEX/CGCCCAGGT/ZEN/GAAGCAGATCTACAA/3IBkFQ/-3' | nt 2370 – 2474 | |
| | mRNA1273-2-R | 5'-AGGATCTGGCTGAAGTTGAAG-3' | | |
| | mRNA1273-1-F | 5'-GACCTTCCTGCTGAAGTACAA-3' | | |
| | mRNA1273-1-P | 5'-/56-FAM/ACCAAGTGC/ZEN/ACCCTGAAGAGCTTC/3IABkFQ/-3' | nt 876 – 1010 | |
| 7 | mRNA1273-1-R | 5'-AAGTTGCTGGTCTGGTAGATG-3' | | nt 876 – 3477 (2602 bp) |
| | Deg-3-F | 5-'GTGACACAGCGGAACTTCTA-3' | | |
| | Deg-3-P | 5'-/56-HEX/TCATCACCA/ZEN/CCGACAACACCTTCG/3IABkFQ/-3' | nt 3367 – 3477 | |
| | Deg-3-R | 5-'GGGTCGTACACGGTATTGTT-3' | | |
| | Deg-2-F | 5-'CACCAACCTGGTGAAGAACA-3' | | |
| | Deg-2-P | 5'-/56-FAM/CGTGCTGAC/ZEN/CGAGAGCAACAAGAA/3IABkFQ/-3' | nt 1647 – 1731 | |
| 8 | Deg-2-R | 5-'CTGCTGAAAGGGCAGGAAT-3' | | nt 1647 – 3477 (1831 bp) |
| | Deg-3-F | 5-'GTGACACAGCGGAACTTCTA-3' | | |
| | Deg-3-P | 5'-/56-HEX/TCATCACCA/ZEN/CCGACAACACCTTCG/3IABkFQ/-3' | nt 3367 – 3477 | |
| | Deg-3-R | 5-'GGGTCGTACACGGTATTGTT-3' | | |
| 9 | mRNA1273-2-F | 5'-GTGGAGCAGGACAAGAACA-3' | nt 2370 – 2474 | nt 2370 – 3477 (1108 bp) |

| | | | | |
|---|---|---|---|---|
| | mRNA1273-2-P | 5'-/56-FAM/CGCCCAGGT/ZEN/GAAGCAGATCTACAA/3IBkFQ/-3' | | |
| | mRNA1273-2-R | 5'-AGGATCTGGCTGAAGTTGAAG-3' | | |
| | Deg-3-F | 5-'GTGACACAGCGGAACTTCTA-3' | nt 3367 – 3477 | |
| | Deg-3-P | 5'-/56-HEX/TCATCACCA/ZEN/CCGACAACACCTTCG/3IABkFQ/-3' | | |
| | Deg-3-R | 5-'GGGTCGTACACGGTATTGTT-3' | | |
| 10 | Deg-1-F | 5-'CATCATCAGAGGCTGGATCTTC-3' | nt 296 – 405 | nt 296 – 3477 (3182 bp) |
| | Deg-1-P | 5'-/56-FAM/CACACTGGA/ZEN/CAGCAAGACCCAGAG/3IABkFQ/-3' | | |
| | Deg-1-R | 5-'GAACTGGAACTCGCACACTT-3' | | |
| | Deg-3-F | 5-'GTGACACAGCGGAACTTCTA-3' | nt 3367 – 3477 | |
| | Deg-3-P | 5'-/56-HEX/TCATCACCA/ZEN/CCGACAACACCTTCG/3IABkFQ/-3' | | |
| | Deg-3-R | 5-'GGGTCGTACACGGTATTGTT-3' | | |

**Table S8.** Optimised MRM parameters for SM-102, ALC-0315, and Dlin-MC3-DMA detection in plasma by mass spectrometry

| Ionizable Lipid | Precursor m/z | Product m/z | Q1 pre-bias | Collision energy | Q3 pre-bias | Dwell time (ms) |
|---|---|---|---|---|---|---|
| SM-102 | 710.6 | 472.4 | -40 | -38 | -23 | 50 |
| | 710.6 | 454.4 | -40 | -45 | -24 | 50 |
| | 710.6 | 318.2 | -28 | -50 | -12 | 50 |
| | 710.6 | 300.2 | -40 | -48 | -20 | 50 |
| ALC-0315 | 767.0 | 694.6 | -20 | -42 | -36 | 100 |
| | 767.0 | 510.4 | -20 | -47 | -26 | 100 |
| | 767.0 | 483.4 | -50 | -41 | -32 | 100 |
| Dlin-MC3-DMA | 643.0 | 132.0 | -40 | -29 | -29 | 100 |
| | 643.0 | 114.0 | -34 | -37 | -26 | 100 |
| | 643.0 | 87.0 | -22 | -55 | -16 | 100 |

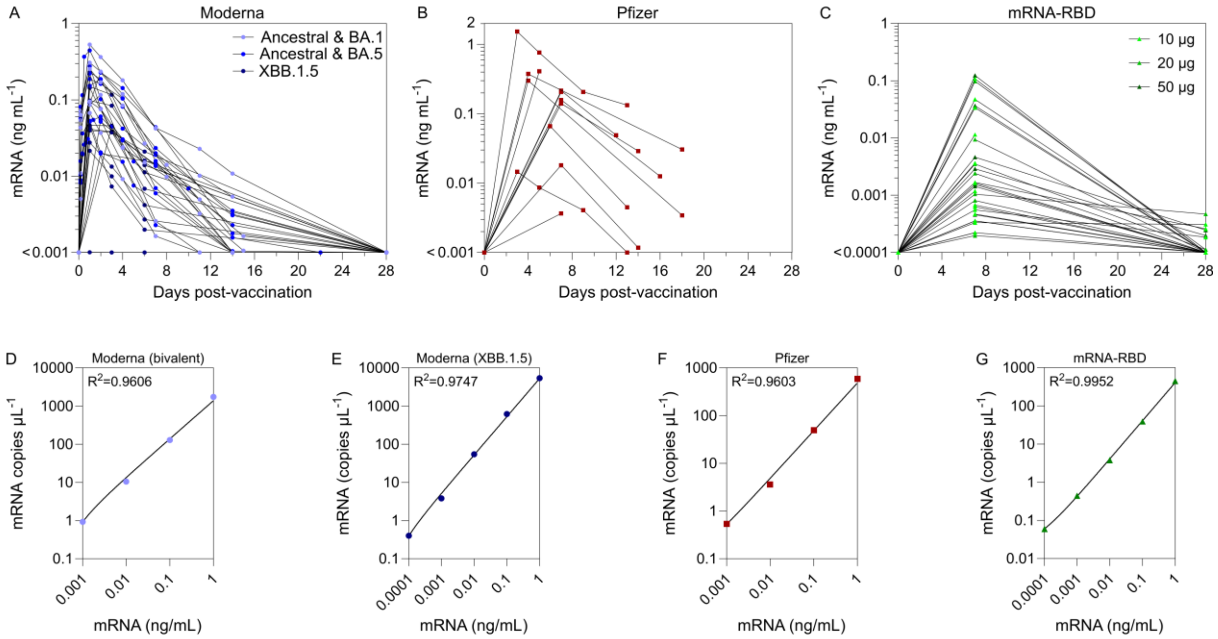


**Figure S1.** Comparison of *in vivo* vaccine mRNA kinetics in human blood following three types of SARS-CoV-2 mRNA vaccination: Moderna, Pfizer, and mRNA-RBD vaccines. (A–C) Longitudinal vaccine mRNA concentrations (ng $mL^{-1}$) in human blood from seven cohorts who received either (A) Moderna bivalent ancestral + BA.1, bivalent ancestral + BA.5, or monovalent XBB.1.5; (B) Pfizer; or (C) mRNA-RBD vaccination at 10, 20, or 50 µg doses. (D–G) Linear PCR standard curves for (D) Moderna bivalent, (E) Moderna XBB.1.5 (F) Pfizer, and (G) mRNA-RBD vaccine mRNA in human plasmas, which were used to determine the lower limit of quantification (LLOQ) shown in Fig. 2 A–C and to covert mRNA concentration from copies $\mu L^{-1}$ to ng $mL^{-1}$. Samples for the linear standard curves were prepared by titrating Moderna bivalent, Moderna XBB.1.5, Pfizer, or mRNA-RBD vaccines (with known mRNA concentration) into human plasma (from one donor collected pre-vaccination) at varying mRNA concentrations.

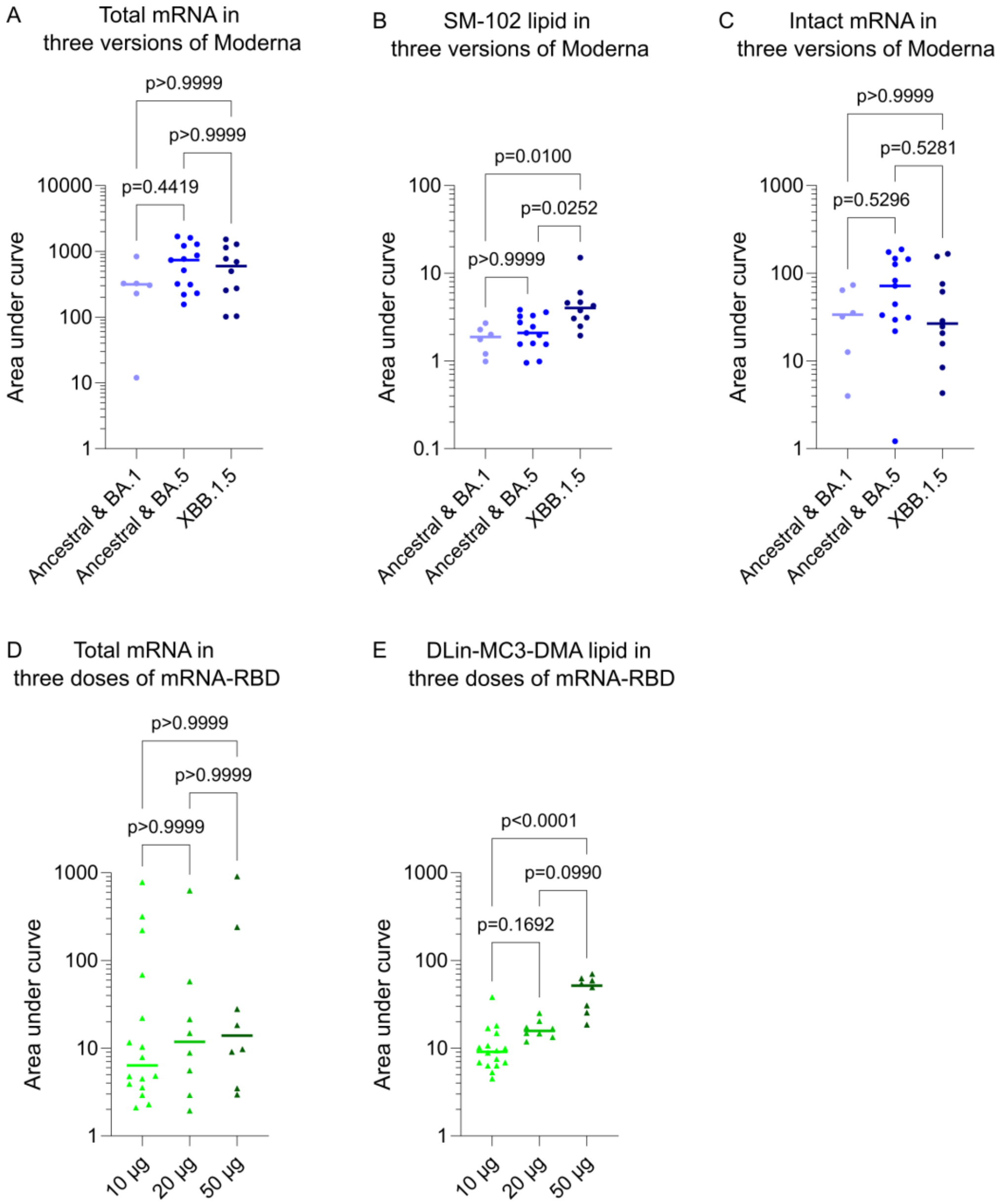


**Figure S2.** Blood distribution of three versions of the Moderna vaccine (bivalent ancestral + BA.1, bivalent ancestral + BA.5, or monovalent XBB.1.5) and three doses of the mRNA-RBD vaccine (10, 20, 50 μg). (A, D) Total mRNA, (B, E) ionizable lipid, and (C) intact mRNA levels in blood were quantified by AUC from the kinetics shown in Figures 2F, 3F and 4E. In (A–E), each dot represents one participant, and the horizontal line indicates the median. Statistical analysis was performed using the nonparametric Kruskal–Wallis test with Dunn's multiple comparisons in (A–E).

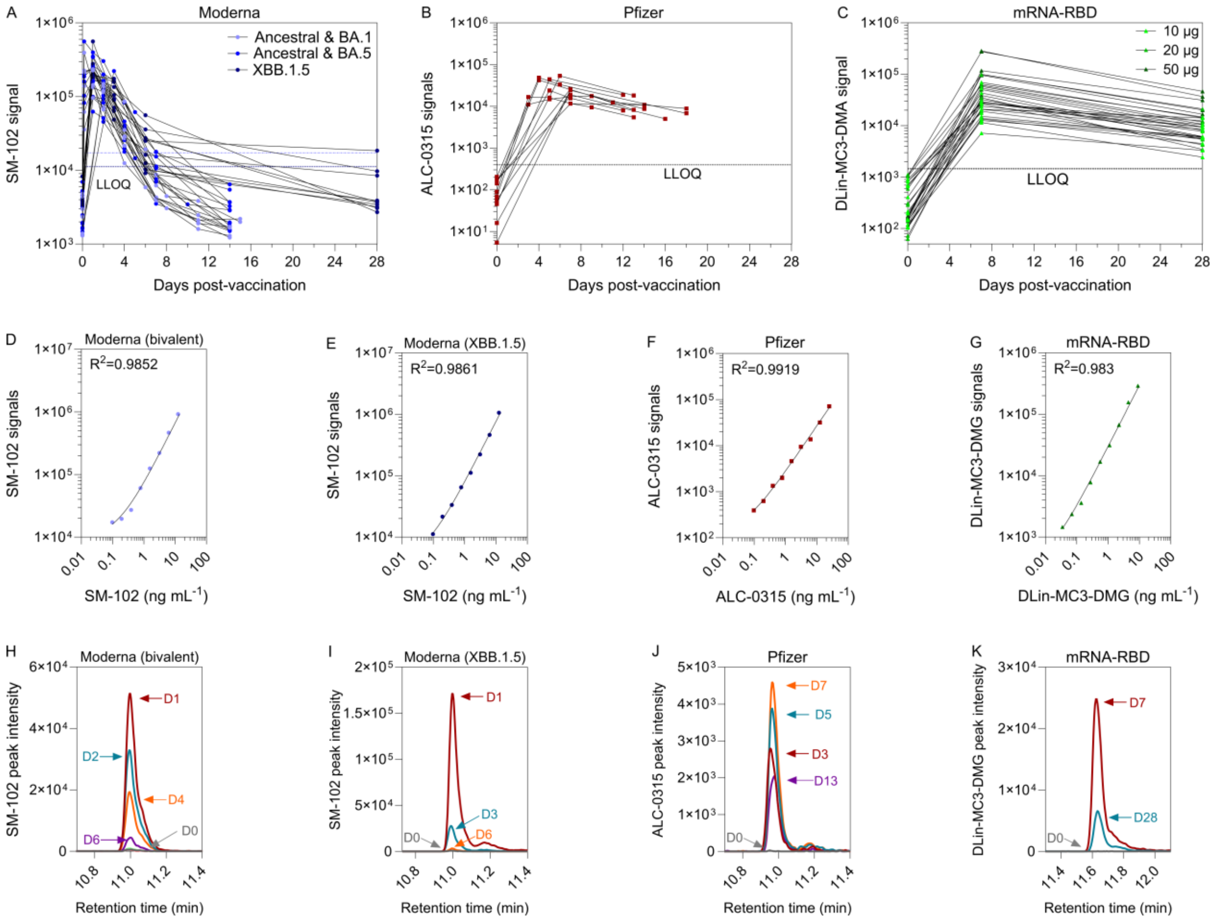


**Figure S3.** Raw mass spectrometry data showing *in vivo* ionizable lipid kinetics in human blood following Moderna, Pfizer, or mRNA-RBD vaccination. (A–C) Longitudinal raw ionizable lipid signals in human blood from seven cohorts who received either (A) Moderna bivalent ancestral + BA.1, bivalent ancestral + BA.5, or monovalent XBB.1.5 (formulated with SM-102); (B) Pfizer (formulated with ALC-0315); or (C) mRNA-RBD (formulated Dlin-MC3-DMA) vaccination at 10, 20, or 50 µg doses. In panel A, two the lower limit of quantifications (LLOQs) are shown: 11207 for Moderna XBB.1.5 (dark blue dashed line) and 17257 for Moderna bivalent vaccines (light blue dashed line). (D–G) Linear standard curves for (D) SM-102 from Moderna bivalent vaccine, (E) SM-102 from Moderna XBB.1.5 vaccine, (F) ALC-0315 from Pfizer vaccine, and (G) Dlin-MC3-DMG from mRNA-RBD vaccine in human plasmas, which were used to determine the LLOQ and to covert raw lipid signals to concentrations (ng mL$^{-1}$) shown in Figure 3A–C. Samples for the linear standard curves were prepared by titrating Moderna bivalent, Moderna XBB.1.5, Pfizer, or mRNA-RBD vaccines (with known ionizable lipid concentration) into human plasma (from one donor collected pre-vaccination) at varying ionizable lipid concentrations. (H–K) Representative chromatograms showing the peak intensity of (H,I) SM-102, (J) ALC-0315, and (K) Dlin-DM3-DMG lipid signals in longitudinal plasma samples from individuals who received (H) Moderna bivalent, (I) Moderna XBB.1.5, (J) Pfizer, or (K) mRNA-RBD vaccination, as determined by liquid chromatography–mass spectrometry (LC–MS).

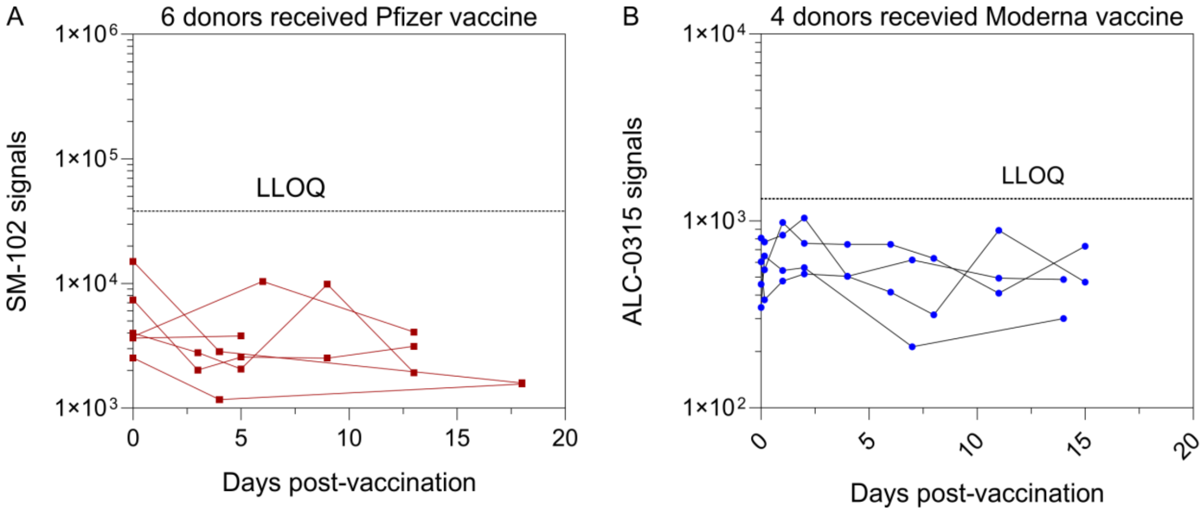


**Figure S4.** Control experiment demonstrating the high specificity of the lipidomics analysis. (A) No SM-102 lipid signals were detected above the LLOQ in longitudinal plasma samples from six donors who received Pfizer vaccination. (B) No ALC-0315 lipid signals were detected above LLOQ in longitudinal plasma samples from four donors who received Moderna vaccination. The LLOQ was 0.1 ng $mL^{-1}$ for both SM-102 and ALC-0315, as determiend from their respective linear standard curves.

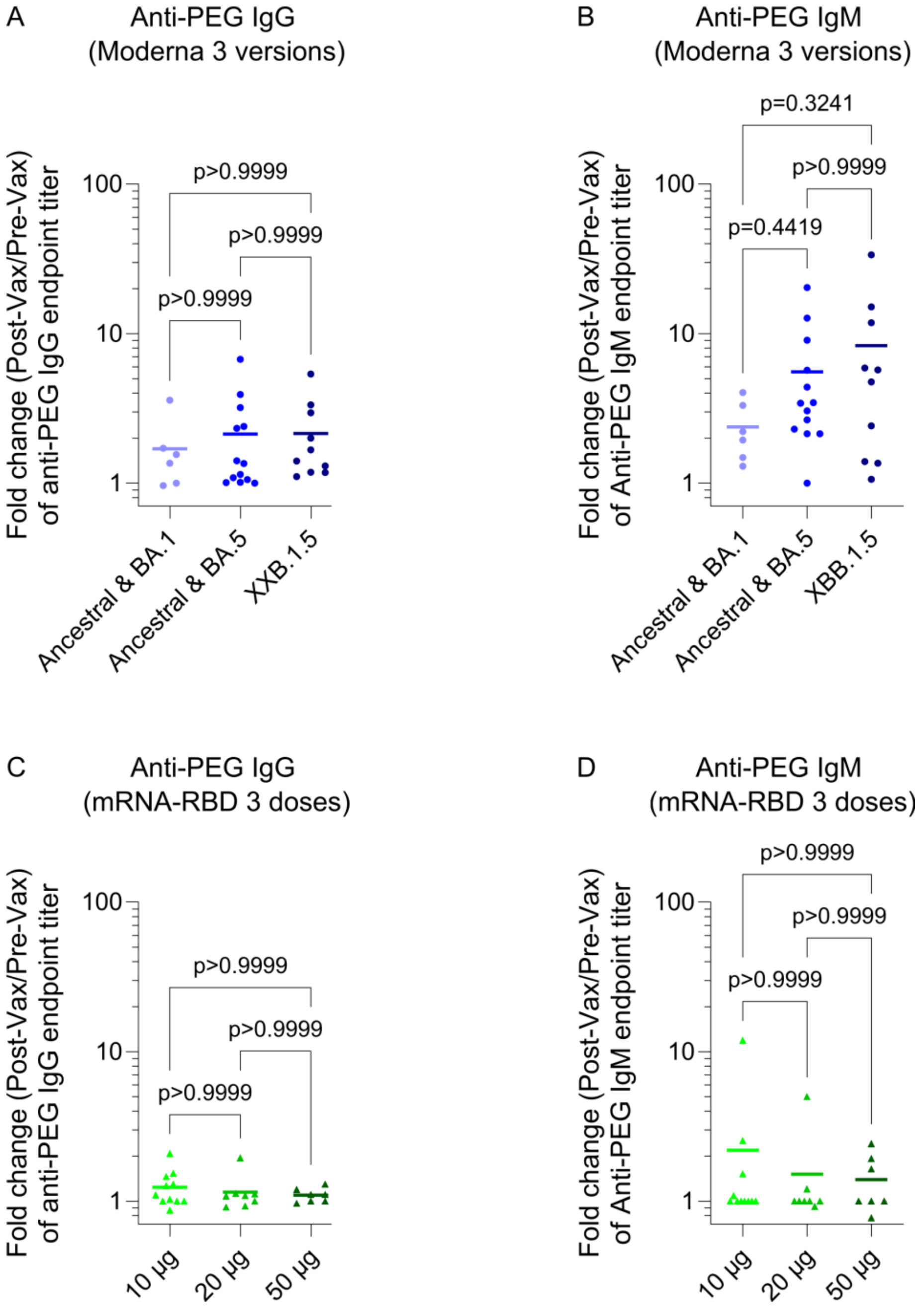


**Figure S5.** Comparison of anti-PEG antibody boosting across three versions of the Moderna vaccine (bivalent ancestral + BA.1, bivalent ancestral + BA.5, and monovalent XBB.1.5) and three doses of the mRNA-RBD vaccine (10, 20, or 50 μg). (A,B) Comparison of fold changes in anti-PEG IgG and IgM titers among the three Moderna vaccine versions. (C,D) Comparison of fold changes in anti-PEG IgG and IgM titers among the three mRNA-RBD vaccine dose groups. In (A–D), each dot represents one participant, and the horizontal line indicates the mean. Statistical analysis was performed using the nonparametric Kruskal–Wallis test with Dunn's multiple comparisons in (A–D).

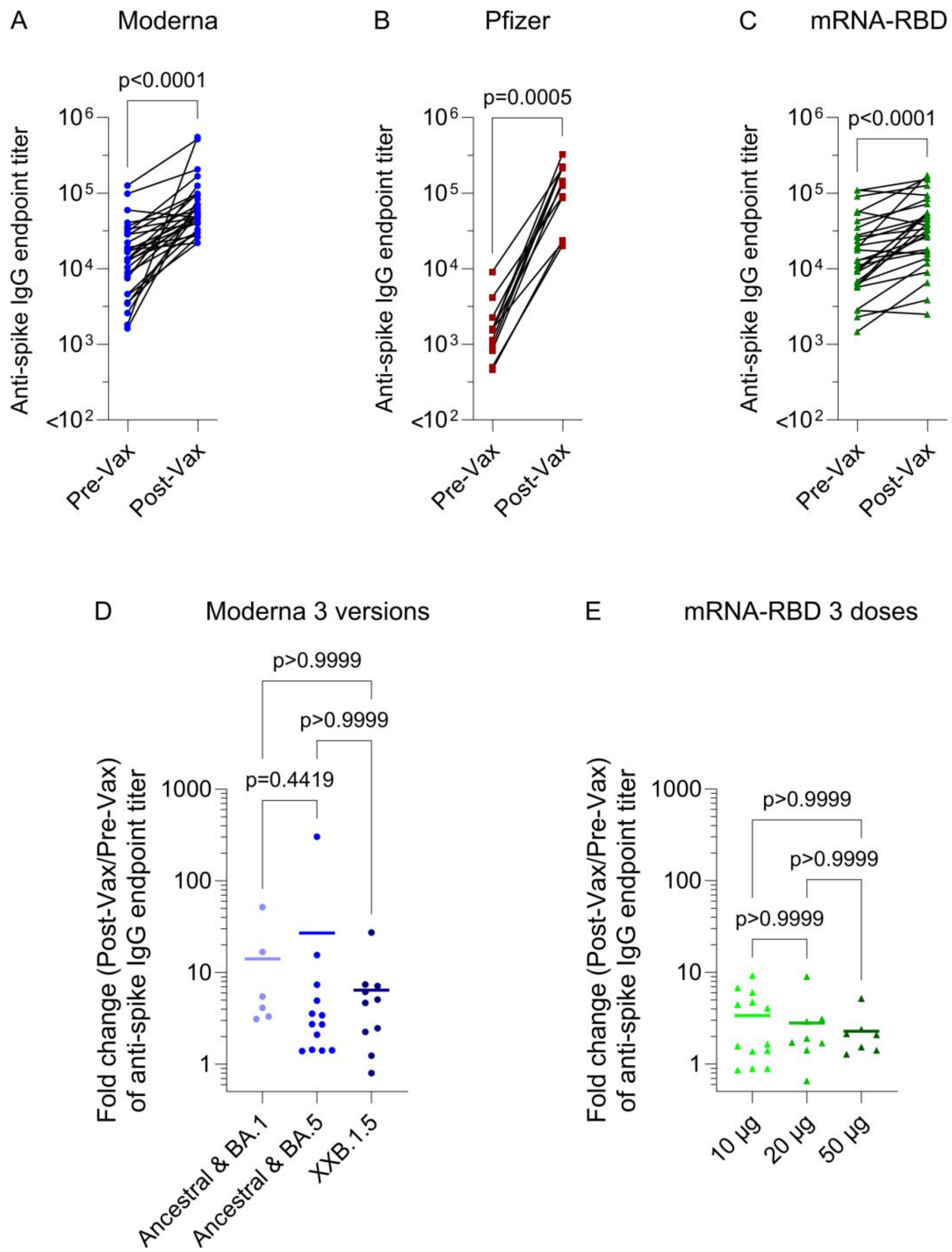


**Figure S6.** Comparison of anti-spike antibody levels in human blood before and after Moderna, Pfizer, or mRNA-RBD vaccination. (A–C) Comparison of plasma anti-spike IgG titers before vaccination (Pre-Vax) and after vaccination (Post-Vax) for the three vaccine platforms. (D) Comparison of fold changes in anti-spike IgG titers among the Moderna vaccine versions. (E) Comparison of fold changes in anti-spike IgG titers among the three mRNA-RBD vaccine dose groups. In (D,E), each dot represents one participant, and the horizontal line indicates the mean. Statistical analysis was performed using the nonparametric Wilcoxon matched-pairs signed rank test in (A–C) and the nonparametric Kruskal–Wallis test with Dunn's multiple comparisons in (D,E).
